# Wearable neuroprosthesis improves mobility and reduces pain in neuropathic participants

**DOI:** 10.1101/2024.05.08.24306164

**Authors:** Noemi Gozzi, Lauren Chee, Ingrid Odermatt, Sanne Kikkert, Greta Preatoni, Giacomo Valle, Nikolai Pfender, Felix Beuschlein, Nicole Wenderoth, Carl Zipser, Stanisa Raspopovic

## Abstract

Peripheral neuropathy (PN) is the most common complication of diabetes. It is characterized by sensory loss which often causes major health consequences including foot ulceration, chronic pain, poor mobility and increased risk of falls. However, present treatments do not counteract the cause of the disease, namely lack of sensory feedback, but rather aim at partial and temporal symptoms relief (e.g. analgesics for pain or creams for ulcers healing). Electrical stimulation is a promising solution for sensory restoration, but it is yet unknown if it can elicit perceivable sensations in PN damaged nerves and whether it could lead to any health or functional benefits. To this aim, we designed a wearable sensory neuroprosthesis providing targeted neurostimulation at the ankle level (NeuroStep) restoring feet lost sensations. We tested it in 14 participants with PN, evaluating its effects on functional outcomes and pain, and the cortical activation related to the restored sensations. Our system was able to restore lost sensations in all participants. The nerves of PN participants resulted significantly less excitable and sensitive than healthy individuals (N=22). Thanks to the neurostimulation, participants improved cadence and functional gait, with even stronger improvements in individuals with higher risk of falls. A full day of NeuroStep use led to a clinically significant reduction of 30.4% ± 9.2% in neuropathic pain. Restored sensations activated cortical patterns, as measured via fMRI, similar to the naturally located foot sensations, thus not requiring training by the user. NeuroStep restores intuitive sensations in PN participants, improving mobility and decreasing pain, possibly replacing multiple inefficient treatments. It holds potential to drastically improve patients’ quality of life thanks to functional and health benefits, while paving the way to new effective neuromodulation treatments.

## INTRODUCTION

Diabetes is a chronic disease affecting 422 million individuals worldwide (*1*). It can occur as a result of genetic or environmental factors and has been linked to obesity and low physical activity (*2*). Diabetes and its complications have a vast financial impact on the healthcare systems currently globally estimated at US$760 billion, with a projected increase to US$825 billion by 2030 (*3*). Diabetic peripheral neuropathy is a neurodegenerative disorder, representing the most prevalent complication of diabetes affecting up to 50% of patients over their lifetime (*4*). Peripheral neuropathy (PN) is slowly progressive (*5*), first affecting long sensory axons causing symptoms such as sensory loss and pain (*6*), followed by autonomic axons, and eventually by larger diameter motor axons (*4*). The mechanisms behind the development and progression of PN are not completely understood but are related to metabolic and micro-vessel alterations which result from chronic hyperglycemia exposure or cardiovascular comorbidities (*7*). Risk factors include glucose management (*8*, *9*), foot care, lifestyle (obesity, smoking, physical activity), and cardiovascular comorbidities (*10–13*). PN can also develop due to many other conditions, as a side effect of chemotherapy, or for unknown reasons (*14*). PN has major health consequences, including foot ulceration, that may lead to lower-limb amputation (*15*, *16*), and neuropathic pain that often leads to depression, anxiety and sleep disorders, as well as reduced quality of life (*17*). Lower-limb complications of PN and neuropathic pain comprise a significant burden for both patients, increasing morbidity and mortality, and for the healthcare system (*16*, *18*). Despite this, treatment options are limited. For instance, commercial medications are available for ulcer treatment, aiming at healing the wound once it is already formed, and avoiding infections that lead to necrosis and amputation. However, they do not address the main mechanism causing the condition, i.e., the lack of sensory feedback and the resulting walking issues and poor balance, thus being unable to prevent the formation of new ulcers. Indeed, the lack of sensory feedback in PN leads to functional deficits during gait as increased risk of falls, and low balance (*19*, *20*) and it is considered the strongest risk factor for diabetic foot ulceration (*12*, *18*). In PN patients, the increased fear of falls, fall-related injuries and the forementioned functional deficits trigger a vicious cycle. Low physical activity and mobility, lead to more progressive diabetes and neuropathy, increased metabolic risks, sensory loss and increased foot ulcerations, which can ultimately lead to limb amputation (*21*). Restoring sensory feedback has the potential to break this vicious cycle, resulting in several health and functional benefits. While sensory feedback restoration has been tested in amputees with both invasive (*22–25*) and non-invasive (*26–32*) approaches, studies in non-amputated neuropathies such as PN are limited. Oddsson et al (*33*, *34*) proposed a solution by providing remapped vibration sensation reflecting plantar pressure measurements at the ankle level. However, vibratory feedback is strongly limited by the bulkiness, rigidity and power consumption of the actuators (*35*) and the remapped approach requires a learning phase to integrate new information during functional tasks, which prevents patients adoption (*29*, *36*). A different approach has been followed by Najafi et al (*21*, *37*), who exploited SENSUS, a stimulator designed for pain treatment, to examine therapeutic effectiveness of plantar electrocutaneous stimulation. They showed some limited improvements in motor-performance and plantar-sensation in participants with PN. However, they do not elicit the distal, somatotopic (lost) sensation, but rather stimulate in-loco (under the electrodes) where the skin area is poorly sensitive and less excitable, while also preventing the dynamic walking of patients. Transcutaneous Electrical Nerve Stimulation (TENS) at the knee level has been adopted by NeuroMetrix QUELL exclusively to treat neuropathic pain (*38*), with limited results (*39*) and without testing its effects on functions and mobility. At the same knee location, TENS showed the potential to restore somatotopic sensations (*30*), projected sensations in the foot sole, but only in amputees and limited to statically laying down (no walking ability).

Considering the limits of knee-positioning, we searched for another leg position, proximal to damaged insensitive feet, where the nerves are sufficiently close to skin to be stimulated via TENS, obtaining the somatotopic sensations in the feet. We hypotheses that this can be achieved close to ankle level. However, TENS has never been tested to restore sensory feedback on damaged nerves as in PN patients, and its feasibility to elicit perceivable sensations and whether such stimulation could lead to health, pain, or functional benefits in daily activities has yet to be demonstrated.

To this aim we designed and implemented a wearable non-invasive neuroprosthesis (NeuroStep) to restore impaired sensations of PN participants. This is achieved by stimulating the damaged nerves at the ankle level with a purposely designed neuroprosthetic sock to provide somatotopic projected fields (PFs) in the different foot locations. We objectively evaluated the characteristics and responses of the PN damaged nerves to the injected charge in terms of perceptual threshold and sensitivity to stimulation via psychosomatic measurements. Then, we tested this neuroprosthesis’s ability to dynamically restore lost foot sensations and improve mobility in functional tasks related to daily life activities. To understand how these artificially restored sensations are processed in the brain, specifically in the primary somatosensory cortex (S1), we used functional MRI (fMRI). We explored whether somatotopic sensations restored through neurostimulation at the proximal nerve level, eliciting PFs in the foot, activate the same S1 areas as when the foot is stimulated directly. Finally, we also assessed reported neuropathic pain after a day of use of the NeuroStep system.

## RESULTS

### Targeted purposely designed TENS partially restores lost sensation in PN participants

We developed NeuroStep (**Fig. 1A**), a wearable neurostimulation system restoring in real-time lost feet sensation in PN participants (N=14). This is achieved by stimulating the damaged nerves at the ankle level with a purposely designed neuroprosthetic sock (**Fig. 1A**) to provide somatotopic projected fields (PFs) in different foot locations (**Fig. 1B3**). The neuroprosthesis is composed of purposely designed matrices of multiple electrodes (**Fig. S1**) to target three lower-limb nerves: peroneal, posterior tibial (medial plantar, lateral plantar and medial calcaneal branches) and sural. We engineered the optimal position, number and dimension of electrodes to be able to successfully target the desired nerves. The electrodes are placed between the malleoli on the dorsal part of the foot for the peroneal array, between the heel and medial malleolus for the posterior tibial array and between the heel and lateral malleolus for the sural array (**Fig. S1**). The number of electrodes and overall dimension of the arrays have been designed considering the average anthropometric measures for each specific key area to in the diverse population of PN participants.

**Fig. 1.**
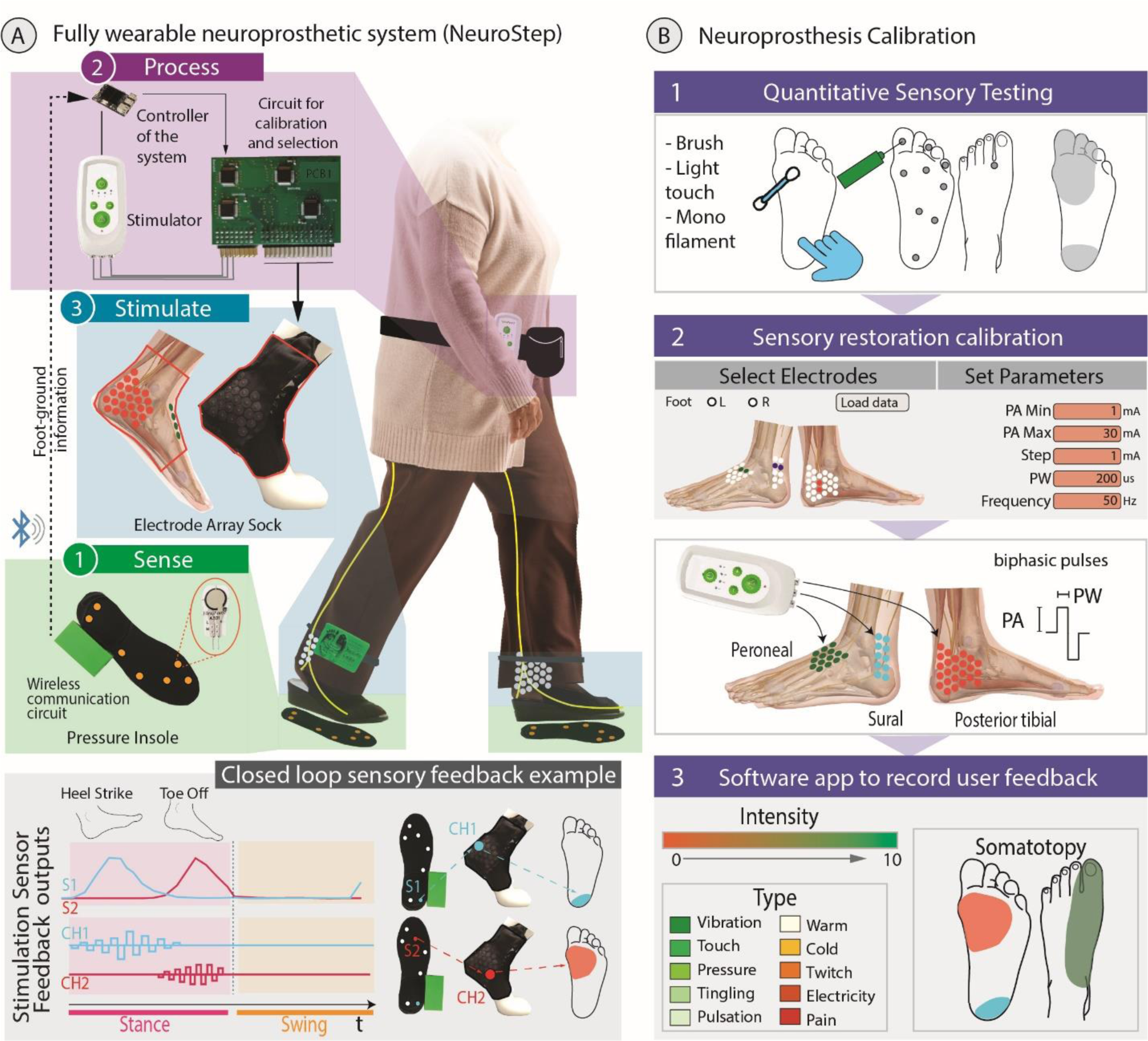
NeuroStep system overview and testing. (**A**) NeuroStep is a wearable neuroprosthesis for closed loop neurostimulation of damaged nerves of PN patients. (1) Force sensitive insole sends foot-ground information. (2) System controller receives insoles data, selects different electrodes and modulates specific stimulation paradigms (found during calibration). (3) Stimulating sock with different electrodes mapped to different insole sensors. During gait, when different sensors are activated, the mapped sock channels are stimulated, and the participant perceives a sensation in the specific areas. (**B**) NeuroStep calibration. 1: Quantitative Sensory Testing was performed with a brush, light touch, and a 10g monofilament to evaluate the sensory loss of PN participants. 2: Electrical stimulation paradigms were calibrated, and appropriate active sites were personalized to achieve targeted sensory restoration. NeuroStep targets three lower-limb nerves: peroneal, posterior tibial (medial plantar, lateral plantar and medial calcaneal branches) and sural. 3: Calibration was performed using a software app to record the user feedback and select the next stimulation paradigm.

For each participant, a personalized calibration phase was performed using a custom designed software app (**Fig. 1B**, **Fig. 2A**), to find the combination of electrodes and electrical charge for an optimal somatotopic and pleasant sensory restoration. The app allows the participant and the experimenters to easily control via software the neuroprosthesis by selecting different electrodes and stimulation paradigm, test them, and directly record the respective perceived sensation (**Fig. 1B**). When the best parameters were found for a participant (combination of electrodes and stimulation that elicit a pleasant somatotopic sensations in the desired areas of sensory loss), these values were saved for dynamical functional experiments. We achieved real time closed loop function (under 50 msec) by mapping sensors in the insoles to different electrodes in the sock. Specifically, the force sensitive insole (1) sends wirelessly foot-ground information to a system controller (2). This controller then activate the specific electrodes of the stimulating sock and modulates personalized stimulation paradigms (found during personalized calibration) (3). During gait, when different sensors are activated (e.g., a sensor on the back of the insole during heel strike and one on the front of the insole during toe off), the specific sock channels that elicit a somatotopic sensation in the pressed area (heel and frontal part of the sole sensations) are stimulated. The accurate design of the NeuroStep neuroprothesis (sensing insoles, processing unit and stimulator with electrodes sock) and its functioning during dynamic walking is shown in **Supp Video S1**.

**Fig. 2.**
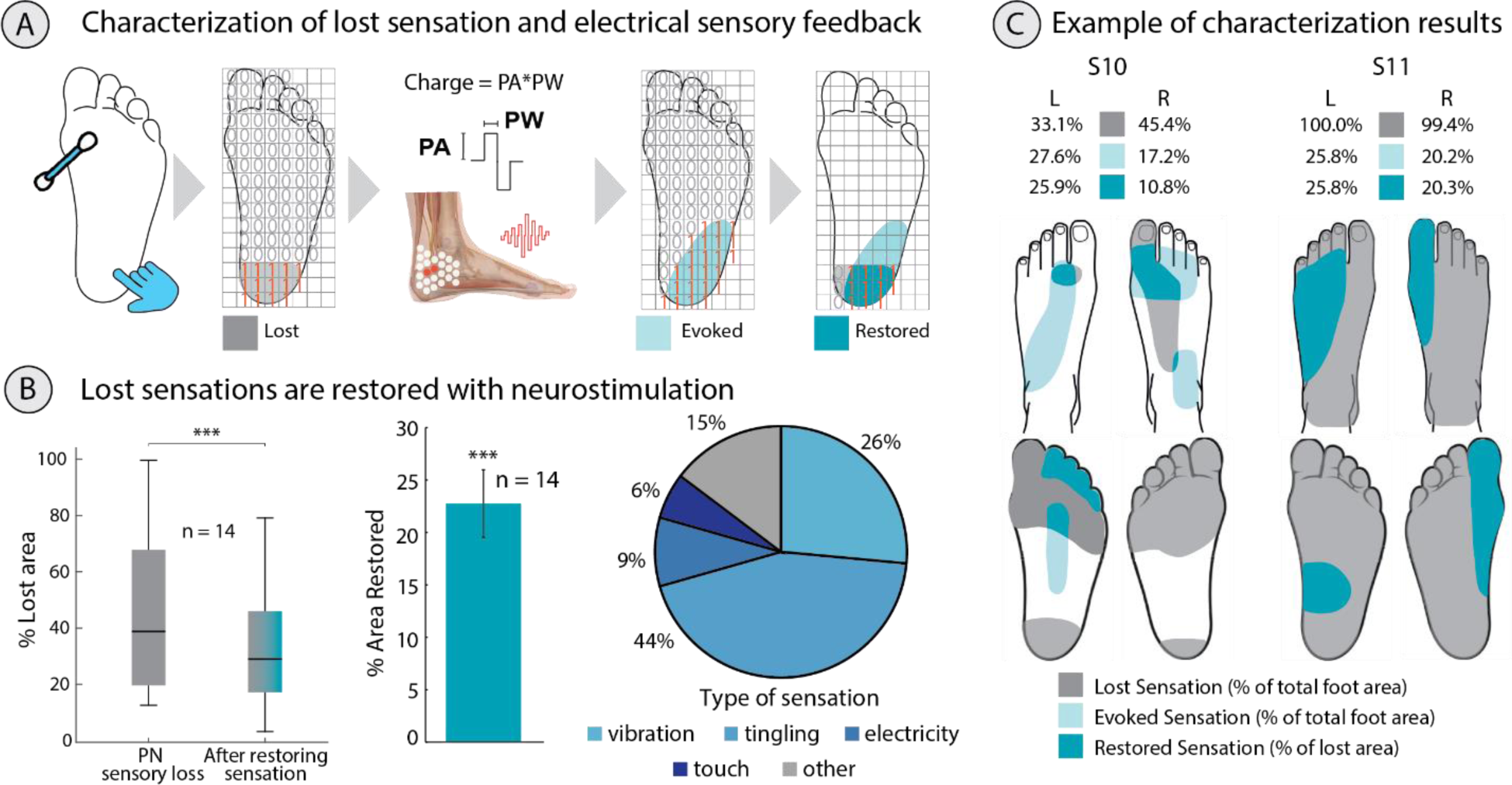
Lost sensations are restored in PN participants. (**A**) Quantification of the perceived area of sensory loss and electrically evoked stimulation using a custom software app with interactive GUI. The areas of sensory loss, elicited and restored sensations are then calculated. (**B**) Lost sensations are restored with targeted neurostimulation. From left to right: decrease in overall sensory loss after restoring sensation (Wilcoxon Signed-Rank test), percentage of lost area that has been restored with neurostimulation (One sample Wilcoxon Signed-Rank test), and type of elicited sensation. (**C**) Example participants with lost, evoked, and restored sensation areas. ***p<0.001.

In **Supp Video S2**, it can be observed that PN participants experienced a loss of touch sensation in various parts of their feet. This is shown through QST on three distinct participants, reporting an absence of sensation when touched while blindfolded on different locations of the sole of the foot. To verify the accuracy of these responses, placebo trials were conducted as well. Quantitative sensory testing (brush, touch and monofilament) (**Fig. 1B, 2A**) showed that PN participants had on average 47.4% area of sensory loss in the left and right feet over the dorsal and plantar areas (**Fig. 2B**). After evaluating sensory loss, we calibrated the neurostimulation to elicit somatotopic sensations overlapping with the sensory loss and we computed the total area of elicited and restored sensations. An example of digitalized feet maps is shown in **Fig. 2C**, with locations of lost, somatotopically evoked and restored sensations. The foot maps and percentages for each participant are shown in **Fig. S2** and **Fig. S3A**. In **Supp Video 2**, we show three PN participants with NeuroStep neurostimulation feedback, able to feel again restored sensations in specific feet locations where previously they reported completely lost sensations. Stimulation successfully restored lost sensations, decreasing the foot sensory loss from 47.4 ± 8.4 to 34.0 ± 6.2 (Wilcoxon Signed-Rank test, p<0.001) (**Fig. 2B**). Overall, we elicited a restored sensation (over the area of sensory loss) of 22.3 ± 3.2% (One sample Wilcoxon Signed Rank test, p<0.001), on the dorsal and plantar sides of the foot (**Fig. 2B**). Different type of sensations, including physiologically plausible ones were elicited (**Fig. 2B**). The most common reported sensations were vibration and tingling (26% and 44% respectively), followed by electricity (9%) and touch (6%).

### PN participants have higher nerve charge threshold and lower sensitivity compared to healthy controls

To quantitatively evaluate sensory loss and capability to restore some of these sensation in PN nerves (**Fig. 3A**), we compared the minimal required charge to elicit a reliable sensation, nerve conduction velocities and sensitivity of PN nerves to healthy controls (N=41) (**Fig. 3B-C-D**). The protocol used to define the charge thresholds similar to the one proposed in (23, 40) can be found in Methods, *Electrical sensory feedback characterization*.

**Fig. 3.**
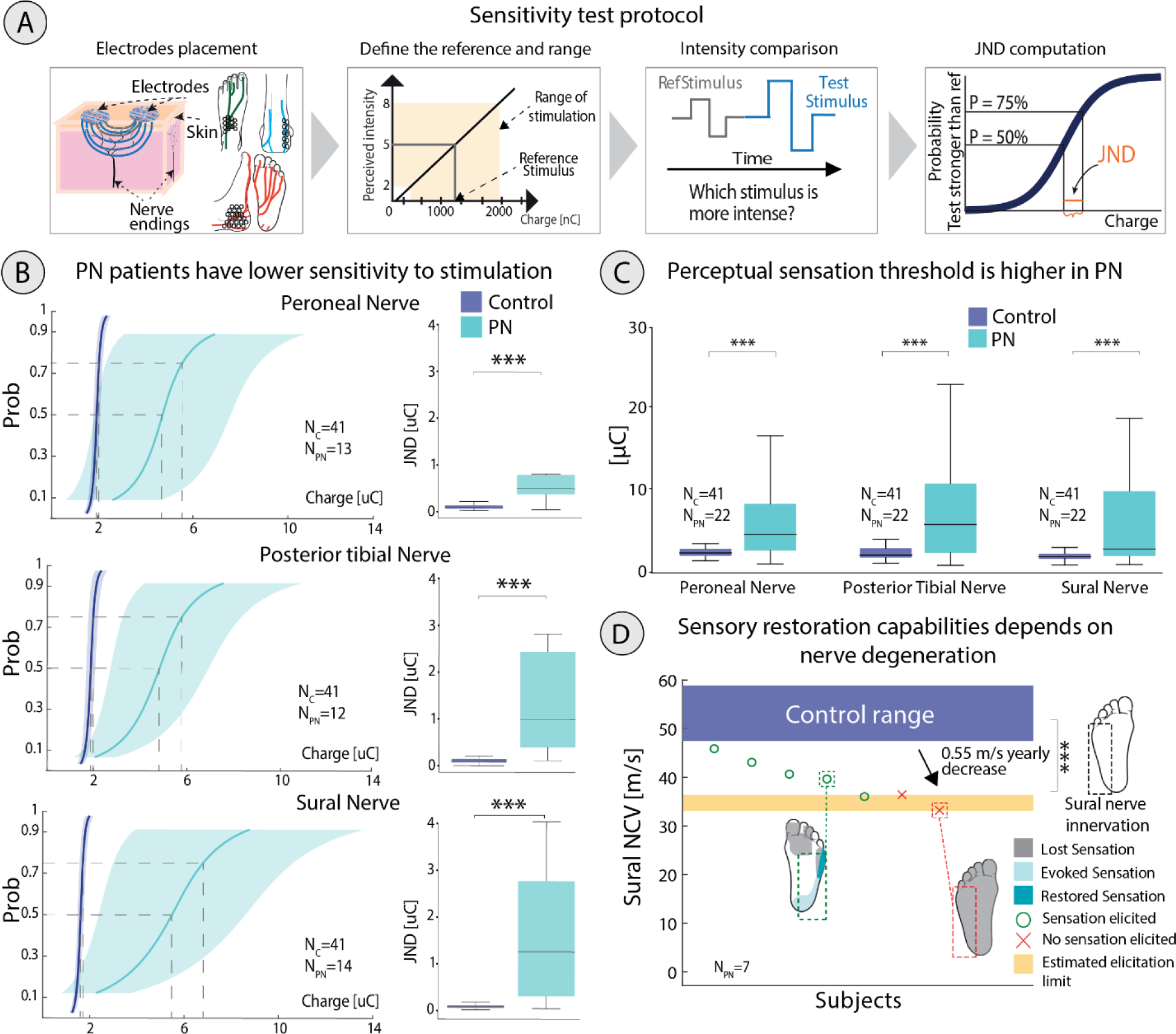
Sensitivity and charge threshold tests for neuropathic nerves vs healthy controls. (**A**) Protocol to test and compute sensitivity through a tactile discrimination task. (**B**) On the left: psychometric curve with standard deviation and boxplots representing the JND of each nerve in PN participants and healthy controls. On the right: comparison of JND (intensity at 75% probability – point of subjective equality at 50% (PSE)) between neuropathic and healthy participants (Mann Whitney U test) (**C**) Threshold charge to elicit a perceptual sensation in neuropathic and healthy control participants (Mann Whitney U test). (**D**) Sensory restoration capabilities depend on nerve degeneration as measured by sural sensory nerve conduction velocity. ***p<0.001.

In PN nerves, the mean perceptual charge threshold to elicit a sensation was 4.1 ± 0.9 µC, 9.6 ± 2.9 µC, and 7.2 ± 1.9 µC in the peroneal, posterior tibial, and sural nerves respectively (**Fig. 3C**). The mean charge threshold in healthy nerves was 1.5 ± 0.1 µC, 1.5 ± 0.1 µC, and 1.2 ± 0.1 µC in the peroneal, posterior tibial, and sural nerves respectively. The threshold charge was significantly higher for PN nerves than healthy controls (Mann Whitney U test peroneal (p<0.001), posterior tibial (p<0.001), and sural nerves (p<0.001)) (**Fig. 3C**). Similarly, the charge required to elicit a strong sensation (i.e., a 8/10 sensation in a scale where 0 is not felt and 10 is unbearably strong sensation) was significantly higher (Mann Whitney U test, p<0.001) for neuropathic nerves than healthy control (**Fig. S4**). We showed that the just noticeable difference (JND), i.e., the discriminability of electrical stimulation (*41*), between PN nerves and healthy controls is significantly higher with neuropathy (Mann Whitney U test, p<0.001). The mean JND in PN was 2377.3 ± 1287.5 nC, 1524.3 ± 475.0 nC, and 2762.0 ± 1164.4 nC in the peroneal, posterior tibial, and sural nerve respectively. The mean JND in healthy controls was 104.1 ± 8.0 nC, 104.5 ± 7.7 nC, and 93.8 ± 6.9 nC in the peroneal, posterior tibial, and sural nerves respectively (**Fig. 3B**). The average Weber Fractions (WF) normalized by reference stimulus (*42*) in each of these nerves are shown in **Fig. S5**. The WFs were significantly higher for PN participants than healthy controls (Mann Whitney U test, p<0.001). Higher WFs indicate a lower sensitivity. The nerve conduction velocities (NCVs) of the neuropathic participants (N=7) measured in the clinical setting with standard sensorimotor nerve conductions tests in the sural nerve were compared to healthy, age matched NCVs reported in the literature (*43*, *44*) and found to be significantly lower (Mann Whitney U test, p<0.001), The mean NCVs of the PN participants included in this study were 39.5 ± 1.0 m/s. The mean NCVs of healthy, age matched controls were 53.2 ± 5.7 m/s (*43*, *44*) (**Fig. 3D**). The sensory NCV (sural) of PN participants was evaluated with respect to the sensations elicited by the neurostimulation in the respective nerve (**Fig. 3D**). NeuroStep was able to restore sensation when NCV was higher than 33 m/s (**Fig. 3D**).

### Walking ability improved in PN participants

Using the previously calibrated electrical sensory feedback and respective charge values, PN participants performed functional tasks (10 m walking test (*45*) and functional gait assessment (FGA) (*46*)) with and without the restored sensation, without any prior gait training. The artificial sensations were mapped to force sensors in the sensorized insole (e.g., sensor under the heel was coupled to a specific combination of electrodes in the sock eliciting a sensation on the heel, Fig. 1A), while the values of the charge were modulated proportionally to the range of force exerted and measured by sensorized insole. One participant did not complete the functional tasks for time reasons. The average FGA score of all participants was significantly improved from 20.7 ± 1.53 without feedback to 23.5 ± 1.16 with feedback (Wilcoxon Signed Rank test, p<0.01) (**Fig. 4A left)**. Of the 13 participants who completed functional gait testing, 9 had a score lower than 23 indicating high risk of falls (*46*, *47*). In this subset of participants, the FGA score improved from 18.6 ± 1.38 without feedback to 22.5 ± 1.36 with feedback (Wilcoxon Signed Rank test, p< 0.001). Single participant results highlighted that 5 individuals, of 9 with a high risk of falls, had a clinically significant improvement in their FGA score (**Fig. 4A right**). Examples of FGA tasks with NF and SF are shown in **Supp Video S3** and **Supp Video S4.** In **Supp Video S3** PN participant S6 significantly improves in two different FGA tasks and explains how the restored sensations through neurostimulation enhanced mobility, coordination and confidence while performing functional tasks. In **Supp Video S4**, PN participants S13 shows a substantial improvement in the balance and speed during a walking task. While initially at risk of falling when using NF, the use of SF enabled S13 to complete the gait task quickly and successfully. The cadence was extracted from the insoles and normalized by walking speed. The median cadence calculated in a 10-meter walk test was found to significantly decrease by 1.16, steps/min with sensory feedback (Repeated Measure ANOVA, p<0.001), i.e., participants were confident in taking longer steps. A significant decrease in cadence was observed in 7 out of 13 participants (**Fig. 4B**).

**Fig. 4.**
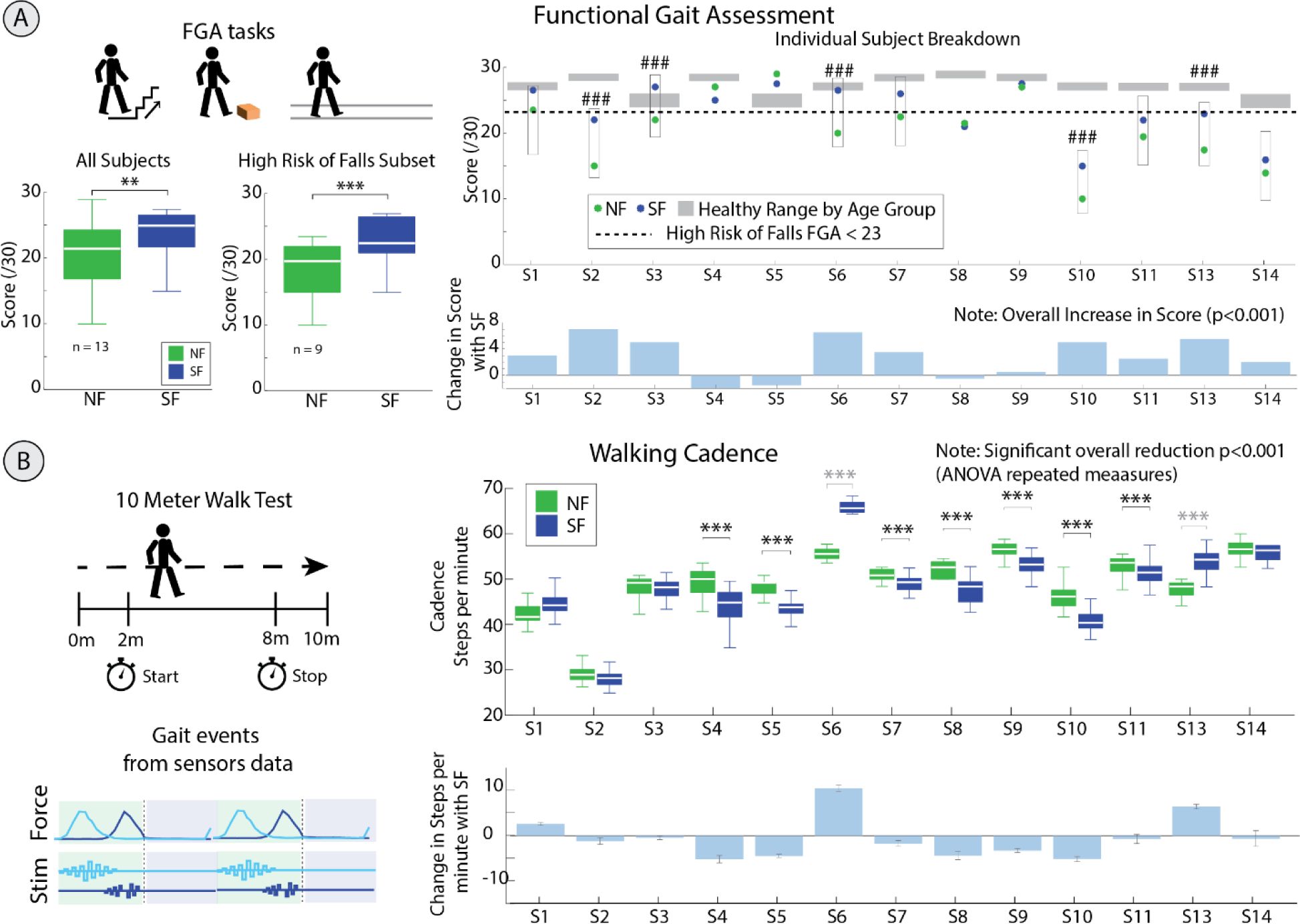
Functional performance improved in PN participants using restored sensory feedback provided by the targeted neurostimulation. (**A**) Functional Gait Analysis (FGA) performance with (SF) and without (NF) sensory feedback. Healthy Age Ranges obtained from Walker et al (*48*). On the left: Box plots show the average score in all participants and in participants at a high risk of falls (Baseline FGA < 23 (*47*)). The individual breakdown shows raw score and change in score. In box plots on the left: Wilcoxon Signed Rank test **p<0.01, ***p<0.001. In individual plots on the right: ### clinically significant change (*33*). (**B**) Walking Cadence with (SF) and without (NF) sensory feedback during the 10-meter walk test. Box plots show the cadence in each participant with SF and NF. Repeated Measure ANOVA *** p<0.001. For single subject results, Wilcoxon Signed Rank test ***p<0.001. S12 did not perform functional tasks and is therefore excluded from this assessment.

### Targeted neurostimulation intervention reduces neuropathic pain

Neuropathic pain was evaluated through the Neuropathic Pain Symptom Inventory questionnaire (NPSI) (*49*) taken before the day of targeted neurostimulation intervention and then again 24 hours later. Cumulative results are shown in (**Fig. 5A)**. Single participant results are shown in (**Fig. S6**). On the day after the intervention, we measured a clinically-significant decrease of 30.4% ± 9.2% of overall NPSI score in the 12 participants who reported pain (**Fig. 5B**) (Wilcoxon Signed Rank test, p<0.05). The dimensions of NPSI mainly responsible for the decrease in pain were burning, paroxysmal pain, and paresthesia, found to clinically decrease by 37.7% ± 11.2%, 48.1% ± 13.3%, and 31.8% ± 14.1% respectively (*50–52*) (**Fig. 5C**). Spontaneous pain and pain attacks were not clinically significant.

**Fig. 5.**
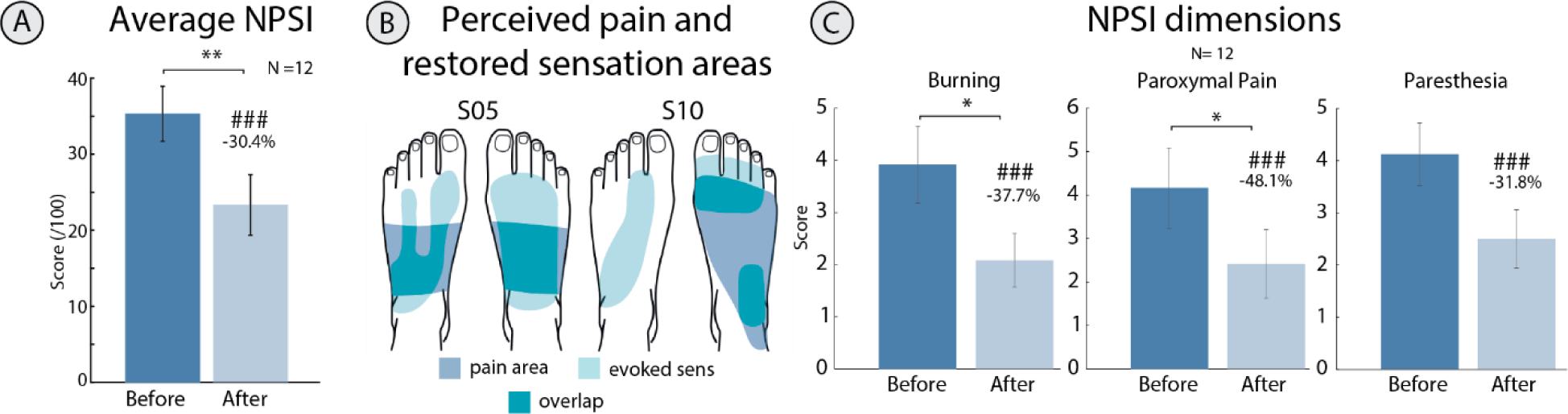
Neuropathic pain symptoms decrease after one day of targeted neurostimulation intervention. (**A**) Average Neuropathic Pain Symptom Inventory (NPSI) before and after intervention. (**B**) Examples of reported areas of pain and evoked sensation. (**C**) Constant pain components of the NPSI. Wilcoxon Signed Rank Test, * p<0.05, ** p<0.01. ###=clinically significant change.

### Somatotopic sensations elicit cortical activity similar to cutaneous foot sensations

Next, we aimed to understand better the neurophysiological basis of the intervention. To this aim, we explored whether restored sensations though targeted proximal stimulation activate the same brain areas as when the foot is stimulated directly. Indeed, due to the somatotopic organization of S1, sensations of each body part (*53–56*) are represented at an anatomically specific cortical location. Healthy (n=12) and PN (n=5) participants underwent 3 Tesla functional MRI, while they were neurostimulated in three different conditions (proximal somatotopic, distal in-foot, control on-ankle, which for simplicity will be called somatotopic, in-foot and on-ankle afterwards) (**Fig. 6A**). More in detail, somatotopic is the targeted proximal nerve stimulation at the ankle level to elicit distal PFs in the foot (as used in the previous experiments). In-foot consists in applying electrocutaneous stimulation locally on top of the previously elicited PFs (naturally located locations over the foot sole). On-ankle is electrocutaneous stimulation at the ankle level avoiding nerve activation, acting as a control condition, where electrodes are placed at a similar location as in the somatotopic condition, but no PFs were elicited. For each of these conditions, three different locations (i.e., corresponding to the peroneal, tibial and sural nerves in somatotopic stimulation) were stimulated (**Fig. 6A and Supp Fig. S7**). Cortical activity evoked by somatotopic neurostimulation were compared to those elicited by in-foot and on-ankle stimulations. We compared overall activity elicited (i.e., across locations) (**Fig. 6B**), spatial similarity across conditions (**Fig. 6C**), and ability to classify (distinguish) the three locations within each condition (peroneal, posterior tibial and sural cortical activation, **Fig. 6D**) in a functionally defined region of interest (ROI) of the S1 foot and ankle area.

**Fig. 6.**
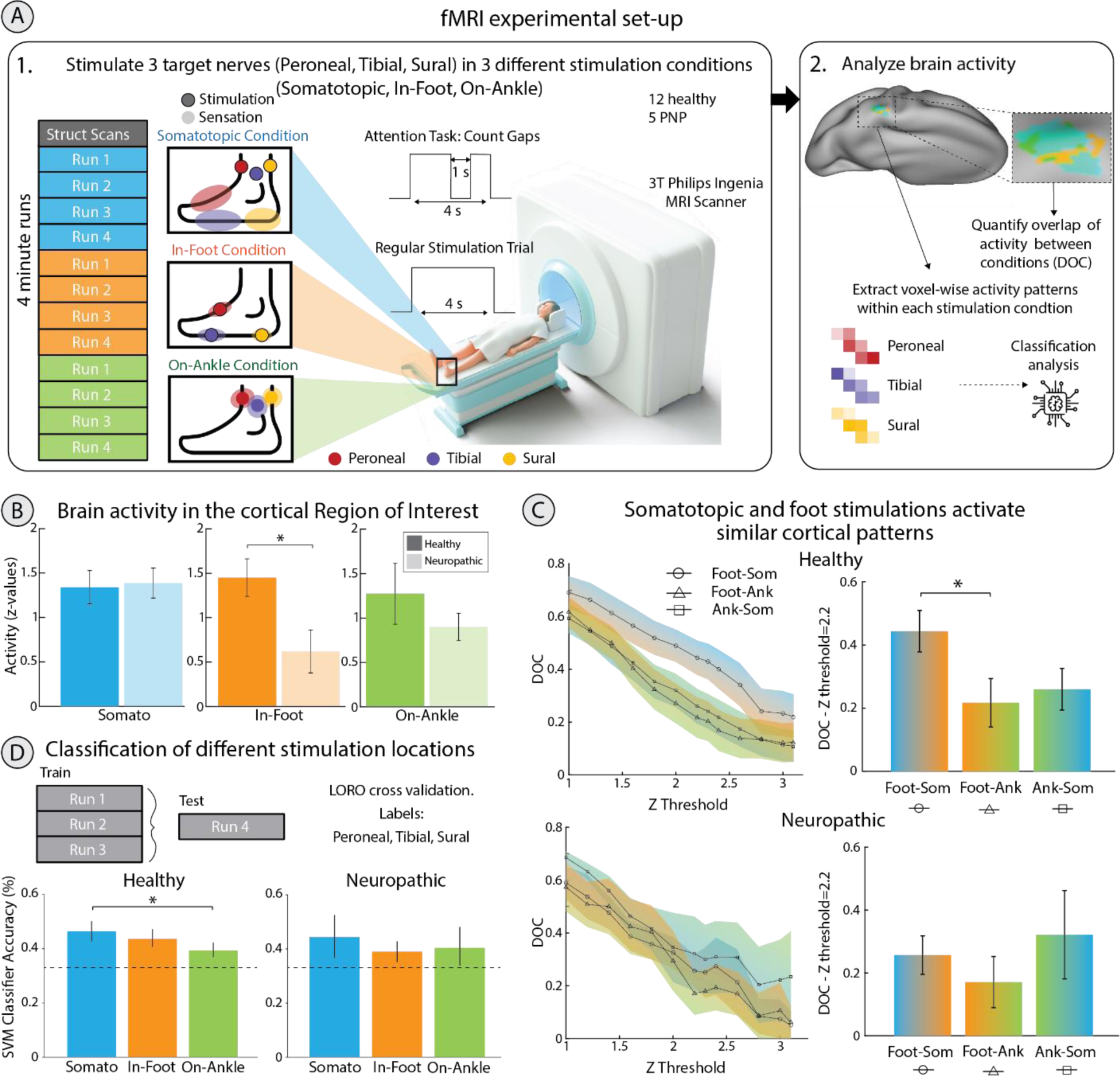
Cortical patterns of somatotopic neurostimulation. (**A**) Experimental Set-Up: The protocol consisted in three different blocks corresponding to the three different conditions: somatotopic, in-foot, on-ankle stimulation. Each block comprised four different runs of 36 stimulations each, in which the three target locations (peroneal, tibial, sural nerves) were presented in a counterbalanced and predefined random order. (**B**) Averaged z-standardized activity in the somatotopic (somato), in-foot and on-ankle conditions in a functionally defined foot and ankle S1 region of interest in healthy (dark colours) and PN (light colours) participants. In-foot activation for PN participants is statistically lower than healthy controls (Mann Whitney test, * p<0.05). (**C**) Spatial overlap of neural activity in the ROI (expressed using the DOC) between in-foot/somatotopic (Foot-Som; circle), in-foot/on-ankle (Foot-Ank; triangle), and on-ankle/somatotopic (Ank-Som; square) conditions. The DOC ranges from 0 (no spatial overlap) to 1 (perfect spatial overlap). The line plots show the DOC for activity thresholded using a Z threshold ranging from 1 to 3.2. Bar plots show the DOC for activity thresholded using Z>2.2. Healthy participants: Friedman test p<0.05, post-hoc Nemenyi test somatotopic/in-foot vs in-foot/on-ankle * p<0.05. PN participants: Friedman test p>0.05. (**D**) Spatial distinguishability of the different stimulation locations (peroneal, tibial and sural) for each condition (somatotopic (somato), in-foot and on-ankle). Average accuracy of a three-class SVM classifier (Leave One Run Out cross validation) among participant is reported. Healthy participants: Friedman test p<0.05, post-hoc somatotopic vs on-ankle * p<0.05.

Resulting cortical activation in the ROI (**Fig. 6B**) during the in-foot condition was significantly lower (Mann Whitney, p<0.05) for PN participants compared to the healthy controls (**Fig. 6B**), as expected, given the reduced or abolished sensations recorded during in-foot stimulation with PN. Instead, somatotopic and on-ankle cortical activation in the ROI were not significantly different between PN and healthy participants (Mann Whitney, p>0.05), indicating that the proximal somatotopic stimulation targets the NP nerves in a location that is still enough healthy to elicit physiologically plausible somatotopic sensations. The spatial overlap between representations of the different conditions within the S1 ROI was computed using the DICE Overlap Coefficient (DOC) (*57*, *58*). The DOC was computed for each condition considering all locations merged. The DOC ranges from 0 (no spatial overlap) to 1 (perfect spatial overlap) and were computed for different z-thresholds. Given the overlap in perceived sensations between the somatotopic and foot conditions, we expected the highest spatial overlap in activity between these conditions and lowest overlap between the foot and ankle conditions. Indeed, we found higher spatial overlap between the somatotopic and in-foot conditions compared to the in-foot vs on-ankle conditions for healthy participants (z-threshold of 2.2, multicomparison with Friedman test p<0.05; post-hoc Nemenyi test somatotopic/in-foot vs in-foot/on-ankle p<0.05, the other post-hoc comparisons were not significant (p>0.05) **Fig. 6C**). Given the low activation of in-foot stimulation for PN subject, we were not expecting a high overlap with somatotopic stimulation. DOC for PN participants was not statistically different among conditions (Friedman test p>0.05).

Finally, we used multivariate pattern analysis to evaluate spatial distinguishability of the three stimulated locations in each condition by applying a SVM classification algorithm with linear kernel. For example, the three nerves stimulated in somatotopic conditions elicit three different foot areas (**Fig. 6A**) which could activate spatially distinct brain patterns in the ROI. In healthy participants, classification accuracies in the three conditions were significantly different (Multicompare Friedman test, p < 0.05). Specifically, the somatotopic had a better spatial distinguishability than the ankle condition (Post Hoc Nemenyi test, p<0.05) which was most likely driven by causing spatially distinct PFs in the foot, while not having the same at the ankle level. Somatotopic vs in-foot and in-foot vs on-ankle were not significant at the post-hoc Nemenyi test. While a trend of higher classification accuracy in the somatotopic condition is observed in the 5 diabetic participants, no significant differences were found between conditions (**Fig. 6D**).

## DISCUSSION

In this study, we engineered and validated a non-invasive wearable sensory neuroprosthesis for restoring lost feet sensation in participants with PN. While after years of disease progressions these participants lost the ability to feel touch in several areas of the foot, we were able to partially restore these sensations. This was possible by using purposely-designed targeted neurostimulation of proximal branches of damaged nerves at the ankle level, where they are still partially healthy, since sensorimotor neuropathies affect the most distal parts of extremities first (*4*, *12*, *59*). However, it was not clear a-priori of the present study whether stimulation of these damaged nerves could elicit distal PFs in the PN feet, within the locations of sensory loss. Achieving that in the totality of patients tested, in at least one nerve, is the first scientific milestone for the viability of the present approach.

However, neuropathy is typically progressive (*5*, *6*), therefore it is important to understand the extent of validity of the present intervention. The success or failure of sensory restoration depends on the severity of the disease, defined by a critical level of demyelination and nerve fibre loss as measured by NCV. We showed that NeuroStep is functional prior to an NCV decrease to ∼33m/s. This threshold was defined considering only the patients with available clinical NCV. Since the time (potentially infinite) for calibration was limited by patient availability, we might have been able to find a somatotopic sensation in patients with a failed nerve characterization by performing a longer and more specific calibration process. Yet, considering this conservative estimate of threshold and the typical slow progression of PN over decades (*5*) (approximate rate of NCV decrease at 0.55m/s per year (*19*, *60–63*)), we estimate that the device could restore sensation for several years (Fig 3D).

We then investigated the specific characteristics of the artificially restored sensation in terms of charge threshold and sensitivity, to understand the phenotype of PN with respect to healthy nerves. We observed that, the required charge for eliciting a sensation was significantly higher for neuropathic participants than for healthy controls. This is consistent with other studies that found the charge threshold (*64*) and maximum tolerable charge (*65*) being significantly higher in PN. Additionally, the sensitivity to electrical stimuli, as measured by JND experiments, was significantly lower for PN participants than for healthy individuals. Lower sensitivity could negatively affect a PN participant’s ability to recognize the intensity of different stimuli, for example associated with the level of pressure of the heel on the floor. This increase in charge threshold and decrease of nerve sensitivity to stimuli is to be expected, due to the degenerative progression of peripheral neuropathy, which evolves through demyelination and loss of nerve fibers (*66*), and is confirmed by a lower nerve conduction velocity.

This neuroprosthesis was able to elicit somatotopic sensations. Regarding the type of sensation, it was able to elicit homologous sensation, i.e., natural sensation of touch (39), only in a limited number of cases. Indeed, most sensations were described as sensations like vibration and tingling.

Near-natural tactile sensation has been achieved only through invasive neural technologies both at the peripheral level (*22*, *25*, *67*), and at the central level (*68–70*). While restoring natural touch information with non-invasive electrical stimulation remains an unresolved challenge (*27*, *29*, *30*, *35*, *71*), these paresthetic sensations can still be subconsciously integrated in the sensory motor scheme(*40*). Despite not eliciting natural sensation, non-invasive stimulation technologies, still have many advantages, namely that they do not require a surgical procedure, avoiding extensive post-surgery care and complications (*35*). Additionally, they are less expensive and do not have biocompatibility issues (*72*), significantly decreasing the risk of infection which is particularly high for diabetic participants who already have reduced wound healing capabilities(*35*, *73*, *74*).

After achieving the first milestone of partial restoration of lost sensations, we aimed to evaluate the possible benefits of this artificial sensory input for functional tasks related to daily activities. FGA improved overall, and in particular it improved in individuals at a high risk of falls (baseline FGA < 23 (*46*, *47*)). This is crucial, considering that, after possible falls, PN participants have an occurrence of injuries 15 times higher than healthy individuals (*75*). Moreover, falls can result in serious injuries (*76*, *77*) requiring medical treatment (*78*), and reducing health and shortening survival (*79*), that can strongly impact the healthcare system (*80*). At the same time, while using NeuroStep, cadence during level-ground walking improved (lower cadence means fewer, longer steps for the same normalized velocity),which is possibly a measurable reflection of higher confidence, and contribute to lower metabolic consumption and lower cardiovascular risk (*31*, *81*).

Very important for clinical application, NeuroStep reduced pain, as measured by the NPSI questionnaire 24 hours after use. This may be explainable through the gate control theory stating that the stimulation of large diameter sensory fibres blocks the signalling of small nociceptive fibres to the brain and therefore decreases pain (*82*). Our intervention with neurostimulation targets the sensory components of pain by stimulating these large diameter sensory fibers (*52*, *83*, *84*), activating the inhibitory mechanism.

Functional and pain improvements are observed immediately with the use of the system, without any previous training. This contrasts with other systems that were tested over a long period of time (*33*, *34*). Immediate improvements are most probably due to the fact that the restored sensations are somatotopic, matching the position where the sensory information was originally expected, thus not requiring any training or learning phase by the user(*35*, *36*). To confirm the hypothesis that somatotopic sensations are processed at the brain level spatially similarly as direct foot sensations, we exploited fMRI. We stimulated healthy participants with intact foot sensations and analyzed the neural correlates of different stimulations.

We showed that somatotopic stimulation elicited cortical activations that spatially overlapped with the ones elicited by foot stimulation, supporting somatotopic stimulation as a valid physiologically accurate alternative to provide foot sensations. As expected, it was not possible to elicit plausible sensations in loco in the foot of PN participants, which is attributed to the distal neuropathy and consequent sensory loss of these participants. The PN participants reported lower or no (consciously) perceptions and exhibited lower cortical activation during foot stimulation. This further supports the use of stimulation on proximal - still partially healthy - PN nerves, to elicit lost distal foot sensations. Furthermore, fMRI results show the limitation of in-loco electrocutaneous approaches like SENSUS (*21*), that stimulates under the sole of the foot, where PN patients did not perceive sensations both at the conscious level (no sensation reported) and unconscious (low fMRI cortical S1 activation) and where it is not possible to stimulate dynamically during walking.

In addition, somatotopic stimulation in healthy controls elicited three distinct brain activity patterns (which can be used as three independent touch information) that could be decoded with higher classification accuracy than the on-ankle condition. This better distinguishability of locations together with the higher overlap between somatotopic/in-foot (physiologically accurate) with respect to in-foot/on-ankle, make NeuroStep stimulation a better alternative than remapped approaches such as those used by Walkasins (*33*). Indeed, re-mapped sensations at the ankle, not only require a further learning step by the user to match the artificial sensations with the original expected input but also elicit cortical activations which are less distinguishable (*36*). Different levels of nerve and mechanoreceptor damage in PN participants make it difficult to predict results from cortical activity of fMRI. This, in combination with a smaller sample size, may explain why results in the PN participants were not consistent.

The somatotopic stimulation we provide allows for more intuitive, physiologically accurate, cognitive integration (*28*, *85*), potentially achieving more benefits in the long term. Step to further improve the naturalness of this artificial sensation could improve its potential to be cognitively integrated. These benefits combined with pain decrease highlight the wide range of functional and health benefits which this wearable and safe neuroprosthesis has the potential to provide.

Limitations of this work include its short-term nature, yet sufficient for the present proof-of-concept. Given the complex experience of pain and the lack of a control condition, we cannot exclude that other mechanisms, including the engagement of the participants, may have played a placebo effect in the decrease of pain. We were also limited in our ability to restore sensation due to the severity of nerve damage. This could be related to the diverse population of neuropathic subjects with different levels of neuropathy and symptoms. Future selection criteria could include NCV to focus on participants who can be better assisted by NeuroStep. Finally, the expert guided calibration required to successfully target nerves that innervate the foot at a proximal level is currently very time intensive and can be automatized in the future to make long term testing more feasible(*86*).

Future work that focuses on a long-term at-home evaluation, with a bigger cohort of patients, would be able to examine more clinical outcomes of use of this device over time. Through this, different functional and health outcomes (improvement in tactile sensitivity, ulcers’ prevention capabilities among others) will be tested together with possible plastic changes at the brain level.

### Conclusions

In this study, we demonstrated the feasibility of restoring lost feet sensations in PN participants affected by a distal sensory loss, using purposely developed neurostimulation. We showed that PN individuals with impaired plantar sensation and high fall risk improved functional outcomes (e.g., gait), while reducing pain, when assisted with somatosensory neuroprostheses. The brain representation of restored sensations highly overlapped with the cortical representations of the corresponding in loco sensations, allowing for fast and effortless integration of the artificial feedback without prior training. This work showcases the potential of using wearable sensory neuroprostheses to tackle functional, health and pain drawbacks of PN and paves the way for further long-term investigations.

## MATERIALS AND METHODS

### Study Design

All PN participants (N=14) participated in one full day of tests which included a morning and afternoon session. The protocol started with the participant filling out an NPSI before any testing began. Then continued with i) Quantitative Sensory Testing (QST) for sensory loss assessment, ii) neurostimulation calibration for sensory restoration iii) Just Noticeable Difference (JND) to test sensory sensitivity and iv) functional tests. Participant-specific information of the testing session is available in **Table S1**. After the one-day intervention, five out of 14 PN participants were recruited for another half day including calibration of somatotopic, in-foot and on-ankle sensations and fMRI testing. The functional assessments consisted of standard tasks performed either with intervention, (i.e. sensory feedback restored with neurostimulation), or without sensory feedback (i.e., with the system turned off). During the control condition the participants were asked to wear the neuroprostheses that was not providing any neurostimulation. The intervention and control condition were presented in a random order determined before each task by the experimenter. The number of control and intervention trials were balanced for each task. The NPSI was taken again 24 hours after the one taken at the beginning of the testing day. Healthy participants performed the full calibration and the JND test for sensory sensitivity (N=22) and the fMRI testing (N=12). Considering that some of the healthy participants perform the JND test and full calibration of both feet, the final number of available nerve data is N=41.

### Participant Recruitment

PN participants were recruited through the University Hospital Zurich and Balgrist University Hospital in Zurich. A total of 14 PN participants were included in the study. The experiments were in accordance with the Declaration of approved by the ETH Zurich’s ethics commission (EK 2019-N-97, EK2021-02258). The trial was registered with ClinicalTrial.gov (NCT05483816 and NCT04217005). All 14 had diagnosed neuropathy with sensory loss and 12 out of the 14 had neuropathic pain. A summary of the participants included in the study is presented in **Table 1** below. Further details are presented in **Table S1**. Sample size (n=13) was computed a priori using an effect size of 1.13 (*21*), α error probability 0.05, 1-β error probability 0.95.

**Table 1.** Recruited Participants.

|  | Age<br>(years) | Sex | Time<br>Since<br>Onset<br>(years) | Diabetes<br>Type | Sensory<br>Loss | Neuropathic<br>Pain |
| --- | --- | --- | --- | --- | --- | --- |
| **S1** | 61-65 | M | 39 | I | Yes | No |
| **S2** | 56-60 | M | 46 | I | Yes | Yes |
| **S3** | 76-80 | M | 22 | II | Yes | Yes |
| **S4** | 56-60 | F | 11 | II | Yes | Yes |
| **S5** | 76-80 | M | 15 | II | Yes | Yes |
| **S6** | 61-65 | F | 22 | II | Yes | Yes |
| **S7** | 56-60 | M | 10 | II | Yes | Yes |
| **S8** | 46-50 | F | 30 | I | Yes | Yes |
| **S9** | 51-55 | M | 1 | II | Yes | No |
| **S10** | 66-70 | M | 4 | II | Yes | Yes |
| **S11** | 61-65 | M | 13 | II | Yes | Yes |
| **S12** | 71-75 | M | 17 | II | Yes | Yes |
| **S13** | 61-65 | M | 40 | II | Yes | Yes |
| **S14** | 71-75 | M | 13 | Not diabetic | Yes | Yes |

Included participants were between 18 and 80 years old and diagnosed with neuropathy, having a loss of sensations from the extremities and neuropathic pain. Participants were able to comfortably walk, sit, and stand without assistance and agreed to comply with the instructions given by experimenters. All participants read and signed the informed consent including the use of identifiable images and access to medical records.

### Participants’ Information

Participants filled out a comprehensive information form including personal data (age, height, weight, shoe size, education level, etc.) and a full description of the location, type, and intensity of neuropathic pain including the Neuropathic Pain Syndrome Inventory (NPSI). Participants’ clinical data such as medical records and clinical test results were also obtained with permission from their doctors. Types of clinical data included, when available, were nerve conduction velocity (NCV) of the sensory sural, motor peroneal, and motor tibial nerves, diabetes onset time (if applicable), HbA1c levels over time. All forms recorded on paper were stored in a locked room and when digitalized, were stored on an encrypted hard drive.

### NeuroStep System

We developed the NeuroStep system, (**Fig. 1**) a wearable neuroprosthesis to deliver non-invasive neurostimulation (**Supp Video 1**). The system was used to provide real-time feedback during functional task performance and is composed of three main components:

1. A Force Sensing Component including:

○ Two Insoles (three fabric layers and one latex layer) embedded with seven force sensors (FlexiForce A301, Tekscan, US) in optimized positions to measure foot-ground interaction information (*32*);
○ Two electronic circuits that send the force information via Bluetooth to a system controller (*32*);
2. A system controller (ODROID-C2, Hardkernel Co., South Korea) that 1) receives the information from the force sensors 2) encodes the information into stimulation parameters that sends to the stimulator 3) selects active electrodes in an array via software and a;
3. A stimulation component including:

○ A CE approved portable stimulator (RehaMove 3, Hasomed, Germany) that provides modulated sensory feedback to the user through neurostimulation.
○ Two custom-made MUX-PCBs that select active electrodes in each array for personalised calibration (Fig. S8);
○ Two smart stimulation socks with three matrices of textile electrodes placed in anatomically plausible positions to target nerves that innervate the foot (peroneal, posterior tibial (medial plantar, lateral plantar and medial calcaneal branches), sural) (Fig. S1);

Additionally, the NeuroStep system includes a custom-made graphical user interface (GUI) which can be used to intuitively interact with the system by selecting the stimulation parameters, controlling the stimulation, and selecting the active electrode(s) in the MUX-PCB and socks. The full calibration can be performed via software without changes at the system. The GUI can also save user feedback such as the perceived sensation location, intensity, and type as described in the section titled “electrical sensory feedback characterization”. Parameters and personalized electrodes can be saved to be used after calibration.

### Sensation Characterization

#### Quantitative Sensory Testing (QST)

Quantitative Sensory Testing was performed on each participant to assess their residual nerve function and understand their areas of sensory loss. The participant was blindfolded while the touch receptors on the sole and dorsum of the foot and around the ankle were stimulated with a gloved finger (light touch), cotton swab (brushing) and a 10mg monofilament. Light touch and brush were tested on the entire area of the foot while monofilament followed the standardized 10-points around the foot (*87*). The participant was then asked to report if and where the touch was felt (**Supp Video 2**). The area of sensory loss (incorrect answers in **Supp Video 2**) was saved using a GUI by drawing the loss area on four different views of each foot. Locations where stimulations were not felt or felt in the wrong location (more than 2mm from the tested area (*88*)) were considered as locations of sensory loss. Then, the area of lost sensation over the dorsal and plantar views of the foot was calculated as a sum of the three different modalities of QST and computed as a percentage with respect to the entire dorsal and plantar foot area (**Fig. 2, Fig. S3**).

#### Electrical sensory feedback characterization

The calibration of neurostimulation parameters in neuroprosthetics is highly participant-specific since the perceived sensation relies on the relative position of the superficial electrodes to the underlying nerve and is sensitive to different nerve conditions (e.g., nerve tissue damage in peripheral neuropathy (*89*, *90*)). Additionally, this may change over time due to neural adaptation (*91*) and/or displacement of the electrodes. To address this, we used a personalized calibration phase to find the optimal electrode positions and stimulation parameters for each participant. The NeuroStep sock was purposely designed to be able to target superficial nerves eliciting three differently located, distal sensations in the foot. Each of the sock’s three arrays was designed considering anthropometric measures (*92*, *93*) to ensure there were enough electrodes (e.g., active sites) to cover the intended target area (**Fig. S1**). Each electrode in the array can be defined by software as disconnected, cathodic, or anodic. In this way, several combinations of electrodes with different locations and dimension can be tested until a proper somatotopic sensation is achieved. During the calibration phase, the experimenters chose the cathode and anode electrodes in the sock using the NeuroStep interactive GUI. Participants were instructed to report the intensity of the stimulation they felt on a scale from 0-10 where 0 is not perceived, 1 is barely perceivable, and 10 is a very strong, not bearable sensation (*31*, *32*). Then, a series of electrical pulse trains increasing in amplitude were delivered until the participant reported a clear, but non-painful sensation with 5/10 intensity. If the electrically elicited sensation was good (somatotopic and, overlapping with the area of sensory loss if possible), the location was selected. If the perceived sensation was mainly under the electrodes or in the wrong location, the experimenters selected different electrodes until an optimal location was achieved. After the electrodes were fixed, a calibration procedure similar to the one described in (*22*, *31*, *32*) was conducted to find the perceptual (minimum, 2/10 intensity) and maximum threshold (below pain, 8/10 intensity) for each nerve for each participant. Since TENS can elicit different types of sensations(*40*), each paradigm was also qualitatively described by choosing sensation descriptors between but not limited to light touch, vibration, tingling, pressure, slow pulsation, twitching, electricity, pain (*94*). Pulse trains consisted of biphasic, rectangular pulses at fixed frequency (50Hz, as previously used in other studies (*29*, *71*, *95*)) while modulating amplitude and pulse width. Each pulse train lasted for 2 s with a 1 s pause. The software automatically increased amplitude or pulse width of consecutive pulses from the minimum to the maximum possible value by a step that is specified by the experimenter. When either the perceptual or maximum threshold was reached, the participant described the perceived sensation in terms of type (vibration, pressure, tingling, warm, electricity, etc), area elicited (dorsal and ventral) and intensity (1–10) in the GUI. For each nerve, the calibration was repeated six times and the charge values were averaged across all trials; the reported sensation area was requested only during the first three trials for each nerve and averaged among them. The electrode position, stimulation parameters and reported sensation were saved for each nerve of each participant using the customized GUI. The area of reported sensation was saved by drawing on four different views of each foot in the GUI. Then, the area of reported sensations was defined as the total area over three different neurostimulation trials for each location/nerve. The total area of artificially evoked sensations was computed as with respect to the entire dorsum, plantar and foot area (Fig. 2D). Then, the area of restored sensation was computed as the intersection between evoked sensation and lost sensation with respect to the entire lost area.

#### Just Noticeable Difference (JND)

The just noticeable difference (JND) is the smallest difference (i.e., the sensitivity) between two stimuli that can be reliably discriminated by the participant (*96*). To determine the JND, two 1-s bursts of stimulation were delivered separated by a break of 1 second (*31*). The range of stimuli was fixed between the perceptual (2/10) and maximum threshold (8/10) found during calibration. The pairs of stimuli consisted of a reference stimulus (average value of the charge range) and test stimulus (a value between perceptual and maximum thresholds). Test stimuli values were computed by modulating the pulse width to find eight equally distributed values between the perceptual and maximum threshold levels around the reference stimulus. Pairs of test stimuli were presented eight times each in a random order with the participant asked to identify whether the first or second stimulus was stronger. This test was performed with 41 healthy participants and 13 neuropathic participants. Depending on time available, each participant did either both or a single foot. One participant did not perform the JND test for limited time available. Then, the probability of judging the test stimulus as larger than the reference stimulus was computed and, a psychometric function was fitted to the probability points (**Fig. 3A**). Then, we computed the point of subjective equality (PSE), i.e., the PW that was perceived higher in at least 50% of the trials and the JND as the difference between the PSE and the PW that was perceived stronger in 75% of the trials (*97*, *98*). For each JND, we computed the WF, which is the JND divided by the reference stimulus. Smaller WFs indicate a higher sensitivity, i.e. a smaller difference in input (PW modulation) is needed to perceive a difference in intensity. Each PN participant performed a different number of JND tests (side and nerves) depending on which nerves were elicited during the calibration and time availability. The overview of JNDs test is summarized in **Supp Table 3**.

### Functional Assessments

The participant was firstly asked to perform few steps (min 10 steps) to measure minimum and maximum force exerted on each pressure sensor. Then, each nerve (stimulation channel) with a successful restored sensation, was matched to the sensor closer to the elicited location. Then, for each pair sensor/stimulation, the perceptual (2/10) and maximum charge (8/10) found during electrical calibration were mapped to the minimum and maximum force exerted on the force insole for functional use (*31*). During functional use the stimulation was modulated between these two values depending on the foot-ground force.

Standard functional tests (the Functional Gait Analysis (FGA) (*46*), and 10 m walking test (*45*)) were performed in both SF and NF conditions. During the NF condition, the participants were wearing the system, but the stimulation was turned off. The FGA is a 10-item test that has been shown to be effective at predicting fall risk in older adults (*46*) and has been recommended as an outcome measure of adults with neurologic conditions (*99*). Each item is scored from 0 to 3 (3 = normal, 2 = mild impairment, 1 = moderate impairment, 0 = severe impairment) with a maximum score of 30. It was performed once per condition per participant in a random order and it was scored independently by two scorers. If scores conflicted (>1 point difference per task), the recorded videos were revisited by a third scorer to determine the correct score. Final scores were computed as the average between the two independent scorers and compared to the average score for each age group (*48*). The 10 m walking test was performed 6 times in each condition in a randomized order. The time from the 2m to the 8m mark was recorded to determine the speed of each trial. The time recorded was averaged between experimenters. Tasks were video recorded and checked by a third experimenter in case of inconsistencies between experimenters (time difference > 0.5 s). During functional assessment, foot-ground force information containing gait events was recorded and saved. Cadence was later extracted from these gait events by counting the number of steps taken between the 2m and 8m mark and dividing by time. All cadence values were normalized by walking speed to compare between feedback and no feedback conditions.

### Neuropathic Pain

The Neuropathic Pain Symptom Inventory (NPSI) (*49*), a new self-questionnaire specifically designed to evaluate the different symptoms of neuropathic pain based on the last 24 hours, was taken at the start of the one-day of intervention and again 24 hours later. NPSI scores were further sub-grouped in spontaneous pain vs pain attacks and in five pain type categories, i.e., burning, evoked, paroxysmal, pressing/deep, paresthesia.

### Cortical Imaging

#### Scanning Procedures

Twelve healthy participants (43 ± 5.53 years old) and five neuropathic participants (72 ± 2.63 years old) (**Table S1**) participated in the functional magnetic resonance imaging (fMRI) investigation of sensation perception. The three distinct target regions or locations innervated by the peroneal, posterior tibial, and sural nerves were characterized and stimulated separately in three main conditions: somatotopic, in-foot, and on-ankle (**Fig. 6A**). Stimulation was applied to one foot. Foot for healthy participants was chosen randomly, while for PN participants it was preferred the foot where the previous characterization achieved a complete characterization (all three nerves). Instead of the custom sock, sticky electrodes were used and were fixated to the participant’s skin using adhesive tape. Stimulation did not elicit noticeable noise artifacts in preliminary fMRI testing. The somatotopic condition was characterized as in the functional calibration described above. Electrodes were placed at the ankle level to elicit distal PFs in the foot. In the in-foot condition, electrodes were placed directly over the location (i.e., the PFs) of the mechanoreceptors where somatotopic sensation was elicited. In the on-ankle conditions, electrodes were placed at the ankle level with the goal of eliciting no distal somatotopic sensation, while still being close to the location of the somatotopically stimulating electrodes. Stimulation intensity was determined as follows: participants were given 2 s of stimulation in increasing amplitude (pulse width and frequency fixed at 300µs and 50Hz) until the stimulation was perceived to be strong but not painful. In six of twelve healthy participants and four PN participants, this corresponded to a numerical intensity rating of 7 or 8/10 and in six of twelve healthy participants and in one PN participant, this corresponded to a numerical intensity rating of 5/10. In neuropathic participants who do not perceive sensation in the foot condition due to their neuropathy, the appropriate foot charge to stimulate with was calculated a priori. A charge ratio between somatotopic and in-foot charge was calculated from the first eight healthy participants’ data (**Fig. S9**) and this was used to determine the appropriate foot condition charge (100). The neuropathic participants were then stimulated at 2 standard deviations above this ratio to confirm their level of loss. Supp Table 2 displays the perceived intensity for the in-foot condition at charge limit in the five participants. Each condition (somatotopic, in-foot, and on-ankle) was targeted in four separate fMRI runs. In each run, the three target locations (correspondent to peroneal, tibial posterior and sural nerves in the somatotopic condition) were stimulated in a counterbalanced order, with a different order of target locations per run. The four runs belonging to the same condition were executed consecutively to minimize the need to switch electrodes. The order of the conditions was balanced across participants to avoid order effects. During an fMRI run, participants saw a white fixation cross via a visual display viewed through a mirror mounted on the head coil. During stimulation, the fixation cross turned red. A stimulation trial lasted 4s with a jittered pause (2-4s) between stimuli for a total of 12 stimuli per location and per run. To ensure stable attention to the stimulation during the fMRI runs, the stimulation was interrupted in a small amount of stimulation trials (2-4 trials per run). In these interrupted trials, stimulation was provided for 2 sec, after which a 1 sec silent period was introduced, followed by another 1 sec of stimulation. Unity 2020.1.13 was used to deliver stimuli and display of the fixation cross. Care was taken to ensure that the interrupted stimulation trials were equally distributed across the stimulation locations within each run. Participants were instructed to count the number of interrupted stimulation trials and verbally reported this at the end of each run. Head motion was minimized using over-ear MRI-safe headphones or padded cushions.

#### MRI acquisition

MRI data was acquired using a Philips 3 tesla Ingenia system (Best, The Netherlands) with a 32-channel head coil. Task-fMRI data was acquired using an echo-planar-imaging (EPI) sequence using the following acquisition parameters: 40 slices, 2.7mm(*101*) resolution, TR: 2500ms, TE: 30ms, flip angle: 85°, SENSE factor: 2. Anatomical T1-weighted images were acquired using the following acquisition parameters: 1mm(*101*) resolution, repetition time (TR): 7.7ms, echo time (TE): 3.6ms, flip angle: 8°.

#### MRI preprocessing

Data collected for individuals of whom we tested the left foot was flipped on the midsagittal plane prior to data preprocessing and analysis. fMRI preprocessing was implemented using FSL v5.0.7 (https://fsl.fmrib.ox.ac.uk/fsl/fslwiki). Cortical surface reconstructions and visualisations were realised using freesurfer v6.0 (https://surfer.nmr.mgh.harvard.edu/) (*102*, *103*) and Connectome Workbench v1.3.2 (https://www.humanconnectome.org/software) (*104*). Common preprocessing steps for fMRI data were applied using FSL’s Expert Analysis Tool (FEAT): motion correction using MCFLIRT(*105*), brain extraction using automated brain extraction tool BET(*106*), spatial smoothing using a 3mm full-width-at-half-maximum (FWHM) Gaussian kernel, and high-pass temporal filtering with a 90s cutoff. Each participant’s functional data was registered to the corresponding T1-weighted image, initially using 6 degrees of freedom and the mutual information cost function, and then optimized using boundary based registration (BBR)(*101*).

#### MRI analysis

##### Univariate analysis

First-level parameter estimates were computed using a voxel-based general linear model (GLM) based on the gamma hemodynamic response function and its temporal derivatives. Time series statistical analysis was carried out using FILM (FMRIB’s Improved Linear Model) with local autocorrelation correction. To reduce noise artefacts, cerebrospinal fluid and white matter scan wise time series were added to the model as nuisance regressors. Data were further assessed for excessive motion and volumes with an estimated absolute mean displacement greater than 1.35mm (half of the functional voxel size) were scrubbed (maximum percentage of volumes scrubbed in a scan = 5.6%). Contrasts were defined per main condition for stimulation versus rest and, additionally, for overall stimulation versus rest (i.e., across conditions). A fixed-effects second-level analysis, as implemented in FSL’s FEAT, was used to average across each main condition’s stimulation runs and across all runs per participant. For optimal coregistration and functional signal alignment across participants’ univariate analysis was performed on the cortical surface. Cortical surface projections were constructed from each participant’s T1-weighted images. The statistical maps results resulting from each individual’s second-level analysis were projected onto the cortical surface using cortical-ribbon mapping and finally to the fs-LR surface template brain. To create a functional ankle-foot S1 area region of interest (ROI) we calculated a whole-brain group average for the healthy participants. We conducted a one-samples t-test for the contrast overall stimulation versus rest (i.e., across conditions) using permutation testing available in PALM (*107*), using 4096 permutations. Activity was thresholded using cluster-based thresholding corrected for multiple comparisons with Z > 2.3. The resulting activity blob at the anatomical ankle and foot area in the primary somatosensory cortex was then extracted as an ankle-foot S1 area ROI. Averaged z-standardized activity was extracted from each participant’s second-level analysis to examine activity elicited by stimulation in each condition in this ankle-foot S1 ROI.

##### Spatial overlap analysis

The DOC overlap coefficient (*108*) was used to quantify spatial overlap between each pair of conditions. The DOC calculates the spatial overlap between two representations relative to the total area of these representations. The DOC ranges from 0 (no spatial overlap) to 1 (perfect spatial overlap). If A and B represent the areas of two representations, then the DOC is expressed as:

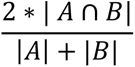

We closely followed previously described procedures(*57*, *58*, *109*, *110*). Activity maps were identified using the contrast stimulation > rest of each participant’s and each condition’s 2nd level analysis (i.e., after averaging across the 4 runs). These activity maps were transformed to the cortical surface (template space), thresholded at Z > 2.2, and masked using both the functional ROI specified above (*111*). To investigate how the choice of threshold influences the spatial overlap results, the DOC was further calculated as a function of z thresholds ranging from 1 to 3 (in steps of 0.2). In-vertex volume between the white matter and pial surfaces was used to calculate the volume covered by the thresholded representations and the resulting DOCs.

##### Classification analysis

To investigate whether the stimulation of the three target locations (peroneal, posterior tibial, and sural nerves) elicits distinct voxel-wise activity patterns in the ankle-foot S1 ROI within each condition, we aimed to decode the stimulated target location from the trial-wise parameter estimates using Multivariate pattern analysis (MVPA). For this, single-trial parameter estimates were computed with an HRF-based first-level GLM in SPM12 (http://www.fil.ion.ucl.ac.uk/spm/) using SPM’s default parameters. The design matrix consisted of individual regressors for each stimulation trial. This resulted in 144 parameter estimates per condition and participant. Within each condition, there were 48 trials per target location. The ankle-foot S1 ROI in the fs-LR template surface space was resampled to each participant’s native 3D space, i.e., the same space as the parameter estimates. Any holes were filled. Next, the voxel-wise parameter estimates below the specified ankle-foot S1 ROI were extracted for each trial and participant.

To investigate whether the stimulation of the three target locations (peroneal, posterior tibial, and sural nerves) elicits distinct activity patterns in the ankle-foot S1 ROI within a condition, we aimed to decode the stimulated target location from the parameter estimates. These extracted 144 trials were used in a machine learning pipeline to classify for each condition the neural activity patterns elicited by the three target locations. Separately for each participant and each condition, a Leave one Run Out (LORO) cross validation was performed. It consists of dividing the dataset by the fMRI run (four) for which data from three runs were used for training and data from the left-out run (i.e., the fourth run) were used for testing. This procedure was repeated four times so that each run served as the testing set once. This ensures independence of the test set from the training set, allowing to prevent overfitting and to generalize performance of the classifier to new and unseen data. At each step, we performed standard scaling, fit on the training set and applied to the test set. The processed dataset was given as input to a SVM classifier with linear kernel. Hyperparameters were set at default values. Models were evaluated in terms of average accuracy on each LORO test set. The average classification accuracies for each condition (somatotopic, in-foot and on-ankle) were compared, separately for healthy controls and neuropathic participants.

### Data collection and statistical analysis

Plotting, data processing and analysis were performed in Matlab (R2022b, The MathWorks, Natick, MA, U.S.A.) and Python 3. Results among patients are presented as average ± standard error. Barplots shows the average and standard error. Boxplots show the interquartile range with median, and the whiskers the rest of the distribution excluded outliers. Statistical analysis was performed using built-in Matlab and Python functions. For clinical measures, published standards for clinically meaningful difference were used to compare baseline and stimulation trials. Non-paired two-groups comparison was performed using Mann-Whitney test (e.g., to compare healthy vs PN participants). Paired comparisons were performed using Wilcoxon Signed-Rank test. Single group comparison against mean was performed using One sample Wilcoxon Signed-Rank test. For functional task comparison at group level, Repeated Measures ANOVA was used. Multicomparison analyses were performed with Friedman Multicompare test and Post-hoc Nemenyi tests. Asterisks on plots indicate the following statistical significance levels: p < 0.05 (*), p < 0.01(**), p < 0.001 (***). ### indicates clinical significance from literature.

## Data Availability

All completely anonymized data, code, and materials used in the analysis are available upon reasonable request to the corresponding author.
Supplementary Videos S2 and S3 are available under request to the corresponding author.

## List of Supplementary Materials

Fig. S1. Electrode Sock Array Design

Fig. S2. Areas of Sensory Loss and Restoration in All Subjects Fig. S3. Restored Sensation by Individual

Fig. S4. Charge Thresholds in Healthy and Neuropathic Participants

Fig. S5. Weber Fraction per Nerve in Control and Neuropathic Participants

Fig. S6. Individual Neuropathic Pain Symptom Inventory Scores Before and After Therapy

Fig. S7. Calibration and areas of sensation of somatotopic, in-foot and on-ankle neurostimulation for fMRI experiments.

Fig. S8. Electrode Selecting MUX-PCB Schematic

Fig. S9. Healthy Charge Ratio for Different Target Nerves

Table S1. Included Subject Characteristics and Task Performance

Table S2. Neuropathic In Foot Sensation Perception

Table S3. JND test for each PN participant

Video S1. NeuroStep System Overview

Video S2. NeuroStep restores missing foot sensations in Diabetic Peripheral Neuropathy Patients. (S2, S3, S5). Patients S2, S3, S5 underwent QST testing showing sensory loss in different areas of the feet. Then, using NeuroStep, we restored the sensations using targeted stimulation and mapped them to specific sensors in the insoles.

Video S3. NeuroStep Improves Feeling and Functionality in Diabetic Peripheral Neuropathy Patient (S6). Patient S6 performs a 10MWT and FGA tests. S6 shows significant improvements in confidence, balance and functionality using NeuroStep sensory feedback.

Video S4. NeuroStep Improves Balance and Speed in Diabetic Peripheral Neuropathy Patient (S13)

## Acknowledgments

The authors are immensely grateful to the volunteers who freely donated their time to the advancement of knowledge and to a better future for traumatic leg amputees. The authors thank Alex Meyer, Aiden Xu, Lara Zamboni for their help in developing the system, Daniel Woolley for writing the Unity program, Manuel Schulthess-Lutz for help with fMRI data collection and preprocessing, and Jana Hajkova for helping in characterization of fMRI experiments. The funder had no role in the experimental design, analysis, or manuscript preparation or submission. All authors had complete access to data. All authors authorized submission of the manuscript, but the final submission decision was made by the corresponding author.

## Author contributions

N.G. and L.C. designed the device, performed all the experiments and all the analyses, made the figures and wrote the manuscript; S.K. and I.O. performed the fMRI experiments and the analyses and wrote the fMRI part of the manuscript; G.P. performed the experiments and prepared the ethical protocol; G.V. co-designed calibration of stimulation; F.B. and N.P. recruited participants; N.W. supervised the fMRI analysis; C.Z. recruited participants, discussed the protocol and results; S.R. designed the device, designed and supervised the experiments, supervised the analyses, discussed the results and wrote the manuscript. All the authors authorized submission of the manuscript, while the final submission decision was taken by the corresponding author.

## Data and materials availability

All completely anonymized data, code, and materials used in the analysis are available upon reasonable request from the corresponding author. Videos S2 and S3 are available under request to the corresponding author.

