## Supplementary Materials for "Wearable neuroprosthesis improves mobility and reduces pain in neuropathic participants"

### Supplementary Material

**Fig. S1. Electrode Sock Array Design**
**Fig. S2. Areas of Sensory Loss and Restoration in All Subjects**
**Fig. S3. Restored Sensation by Individual**
**Fig. S4. Charge Thresholds in Healthy and Neuropathic Participants**
**Fig. S5. Weber Fraction per Nerve in Control and Neuropathic Participants**
**Fig. S6. Individual Neuropathic Pain Symptom Inventory Scores Before and After Therapy**
**Fig. S7. Calibration and areas of sensation of somatotopic, in-foot and on-ankle neurostimulation for fMRI experiments.**
**Fig. S8. Electrode Selecting MUX-PCB Schematic**
**Fig. S9. Healthy Charge Ratio for Different Target Nerves**
**Table S1. Included Subject Characteristics and Task Performance**
**Table S2. Neuropathic In Foot Sensation Perception**
**Table S3. JND test for each PN participant**
**Video S1. NeuroStep System Overview**
**Video S2. NeuroStep restores missing foot sensations in Diabetic Peripheral Neuropathy Patients. (S2, S3, S5)**
**Video S3. NeuroStep Improves Feeling and Functionality in Diabetic Peripheral Neuropathy Patient (S6)**
**Video S4. NeuroStep Improves Balance and Speed in Diabetic Peripheral Neuropathy Patient (S13)**

**Table S1. Included Subject Characteristics and Task Performance.**

| | Sensory Loss | Neuropathic Pain | Just Noticeable Difference | Functional Tests | fMRI |
|---|---|---|---|---|---|
| S1 | ✓ | × | ✓ | ✓ | × |
| S2 | ✓ | ✓ | ✓ | ✓ | × |
| S3 | ✓ | ✓ | ✓ | ✓ | ✓ |
| S4 | ✓ | ✓ | ✓ | ✓ | × |
| S5 | ✓ | ✓ | ✓ | ✓ | ✓ |
| S6 | ✓ | ✓ | ✓ | ✓ | ✓ |
| S7 | ✓ | ✓ | ✓ | ✓ | × |
| S8 | ✓ | ✓ | ✓ | ✓ | × |
| S9 | ✓ | × | ✓ | ✓ | × |
| S10 | ✓ | ✓ | ✓ | ✓ | ✓ |
| S11 | ✓ | ✓ | ✓ | ✓ | × |
| S12 | ✓ | ✓ | ✓ | × | × |
| S13 | ✓ | ✓ | ✓ | ✓ | × |
| S14 | ✓ | ✓ | × | ✓ | ✓ |

**Table S2. Neuropathic In Foot Sensation Perception.** Perceived intensity in loco in the foot reported as an intensity from 0-10 where 0 is not perceived, 1 is barely perceivable, and 10 is a very strong, not bearable sensation.

| | Peroneal Nerve | | Posterior Tibial Nerve | | Sural Nerve | |
|---|---|---|---|---|---|---|
| | Perceived Intensity | Intensity at Stimulator Limit | Perceived Intensity | Intensity at Stimulator Limit | Perceived Intensity | Intensity at Stimulator Limit |
| **S03** | **4/10** | * | **7/10** | * | **0/10** | 0/10 |
| **S05** | **4/10** | * | **0/10** | 2/10 | **0/10** | 1/10 |
| **S06** | **4/10** | * | **2/10** | 4/10 | **2/10** | 2/10 |
| **S10** | **2/10** | 5/10 | **2/10** | 8/10 | **8/10** | * |
| **S14** | **5/10** | 8/10 | **8/10** | * | **4/10** | 7/10 |

* Perceived intensity at stimulator limit not tested

**Table S3. JND test for each PN participant**. Different sides and nerves were tested for each PN participant. First, depending on the characterization, not all nerves were elicited. Then, considering the nerves were a characterization was positive and the limited amount of time for each participant, a random selection of nerves and foot was performed. V: JND test performed. X: JND test not performed. V/X: JND test performed but failure to fit the psychometric curve to the data.

| PN participant | Left – Peroneal | Left – Tibial | Left – sural | Right – Peroneal | Right – Tibial | Right – sural |
|---|---|---|---|---|---|---|
| **S1** | V | V | V | V | V | V |

| | | | | | | |
|---|---|---|---|---|---|---|
| **S2** | X | X | X | X | V | V |
| **S3** | V | V | V | V | V | V |
| **S4** | V | V | V | X | X | X |
| **S5** | V | X | V/X | X | V | V |
| **S6** | V | V | V | V | V | V |
| **S7** | V/X | X | X | X | X | X |
| **S8** | X | V | X | X | X | X |
| **S9** | V | V | X | V | V | V |
| **S10** | X | X | V | V | X | X |
| **S11** | X | X | V | X | X | V |
| **S12** | V/X | X | X | V | X | X |
| **S13** | V | X | V | X | X | X |
| **S14** | X | X | X | X | X | X |
| **Total** | 7 | 6 | 7 | 6 | 6 | 7 |

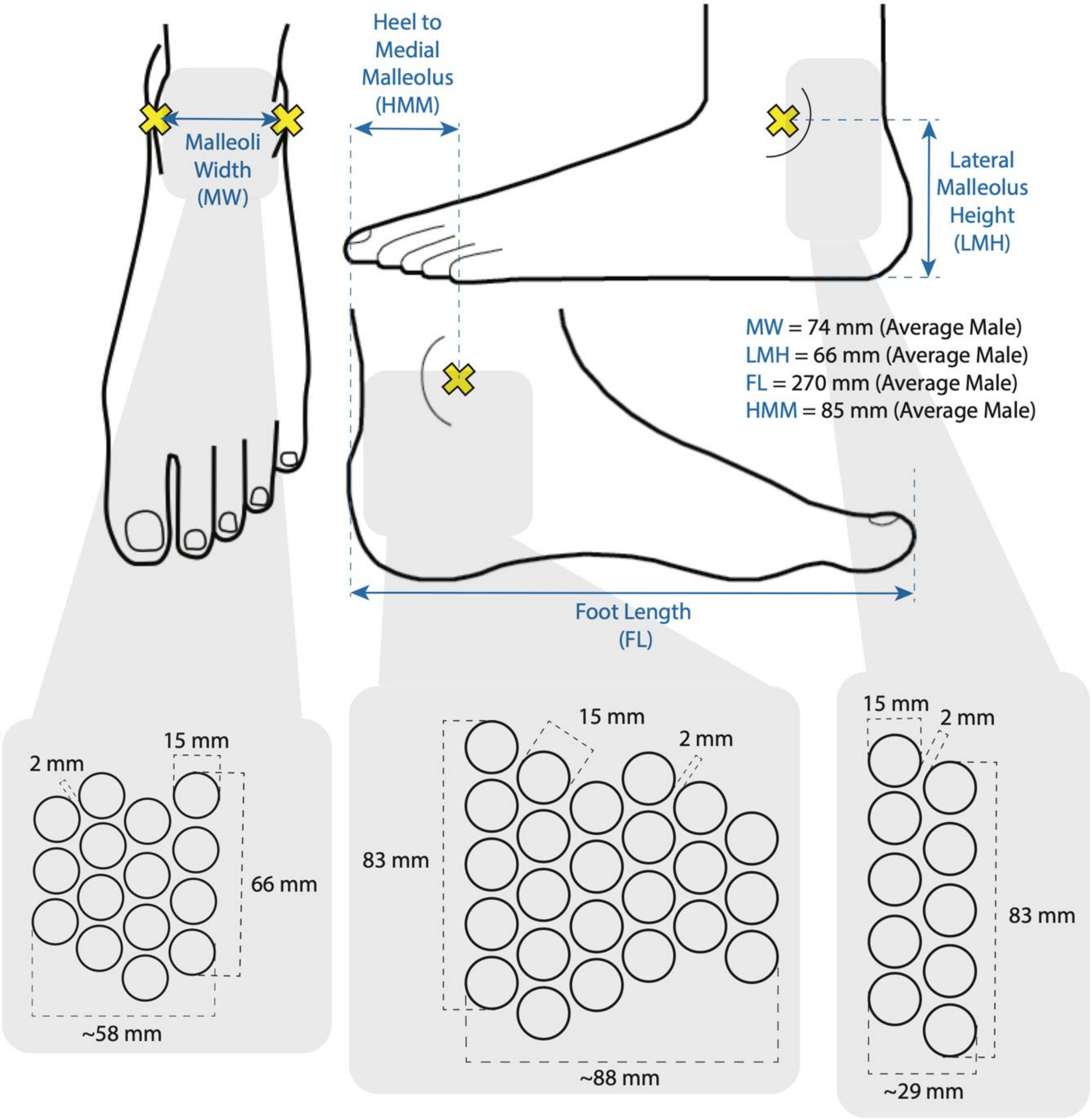


**Fig. S1. Electrode Sock Array Design.** Anthropometric measures for key areas of the foot and ankle and specifications for array design.

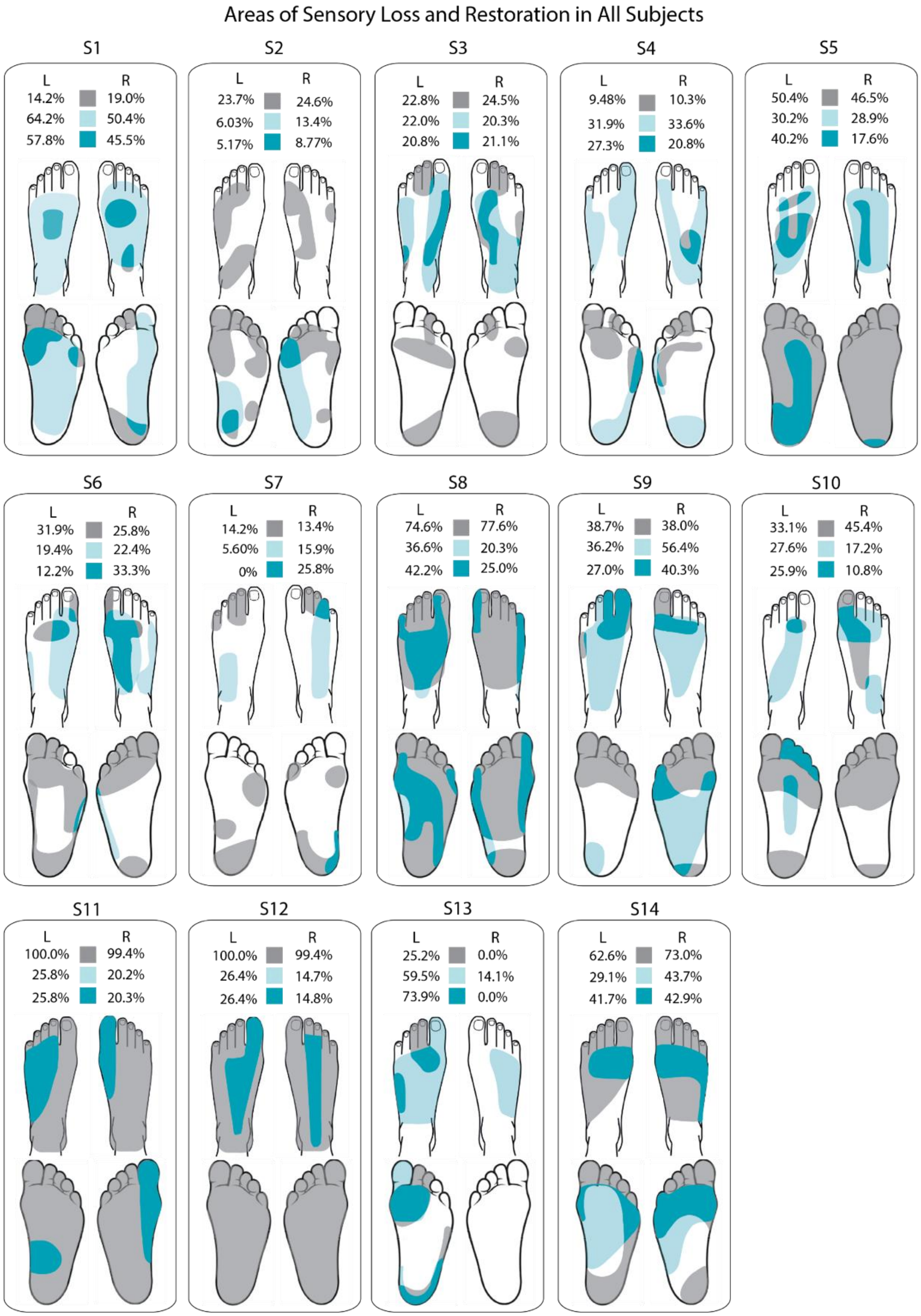


**Fig. S2. Areas of Sensory Loss and Restoration in All Subjects.** Percentage of lost sensation area as evaluated through quantitative sensory testing in grey. Percentage of electrically evoked sensation area in turquoise. Evoked sensation that overlaps with lost sensation is labelled restored sensation.

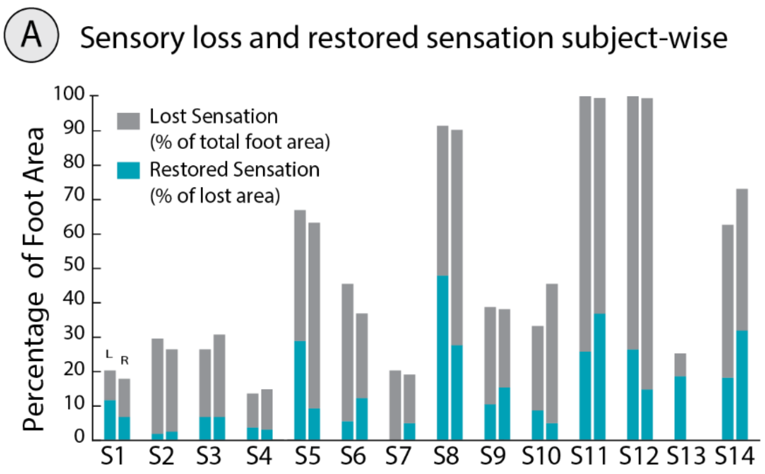


**Fig. S3. Restored Sensation by Individual** (A) Lost and restored sensation on each foot by individual

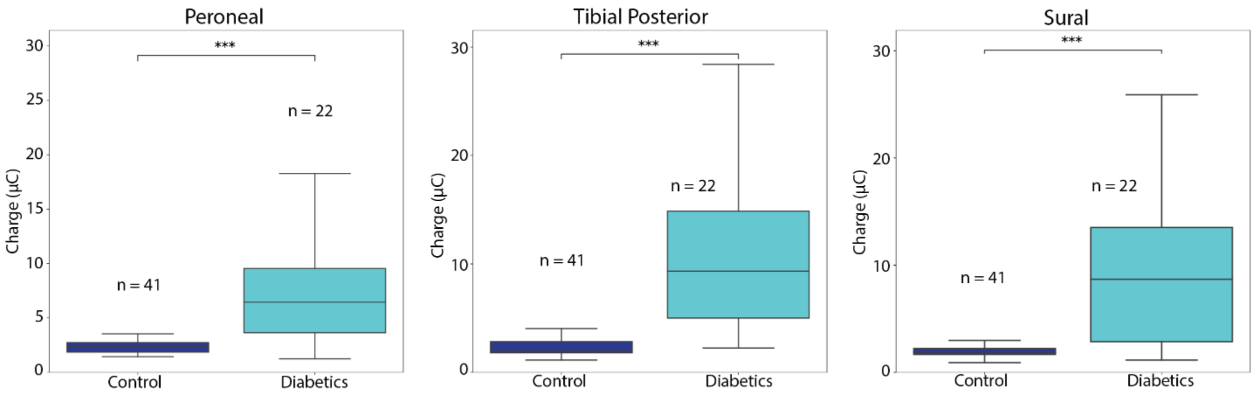


**Fig. S4. Maximum charge in Healthy and Neuropathic Participants.** The charge required to elicit a strong sensation, i.e., a 8/10 sensation in a scale where 0 is not felt and 10 is unbearably strong sensation (MannWhitney U test). ***p<0.001

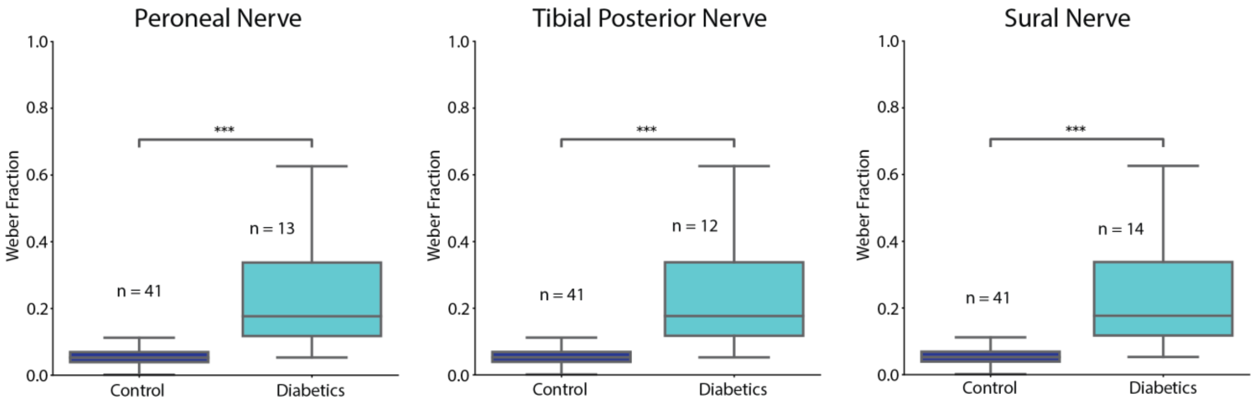


**Fig. S5. Weber Fraction per Nerve in Control and Neuropathic Participants.** Weber fraction calculated as the ratio of JND to reference charge. A Mann-Whitney test was used to compare the distributions. ***p<0.001

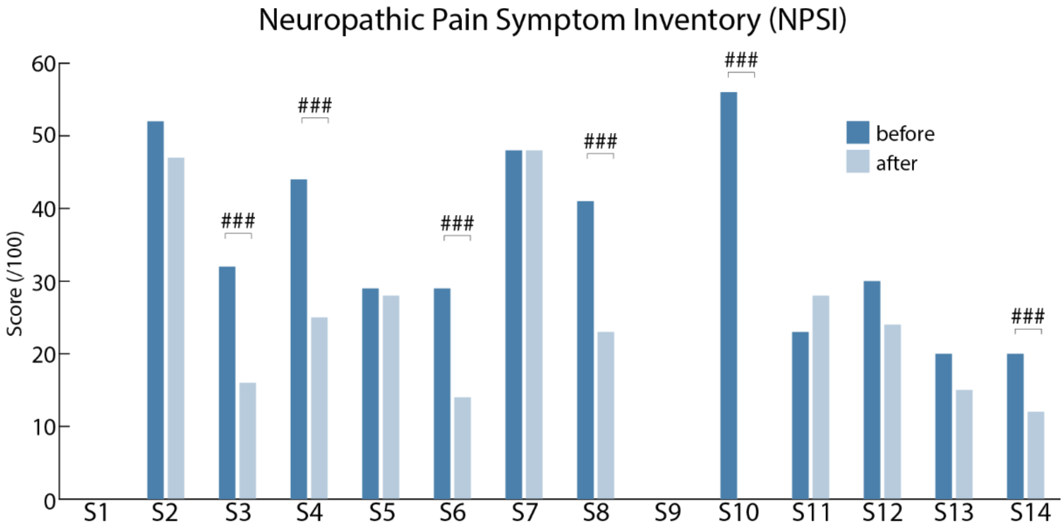


**Fig. S6. Neuropathic Pain Symptom Inventory.** NPSI Scores before and after therapy by subject. ###=clinical significance

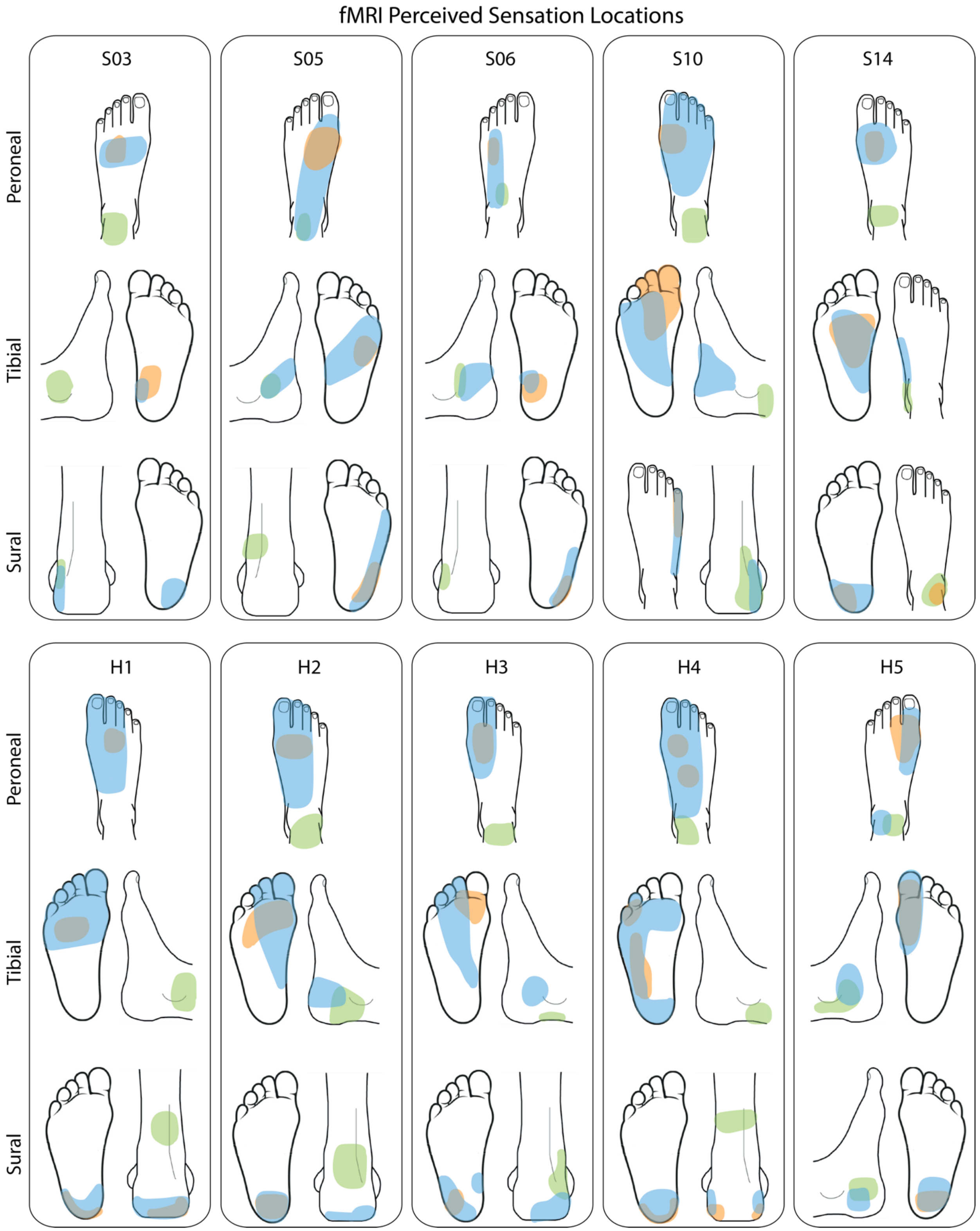

fMRI Perceived Sensation Locations
S03
S05
S06
S10
S14
H1
H2
H3
H4
H5
Peroneal
Tibial
Sural
Peroneal
Tibial
Sural

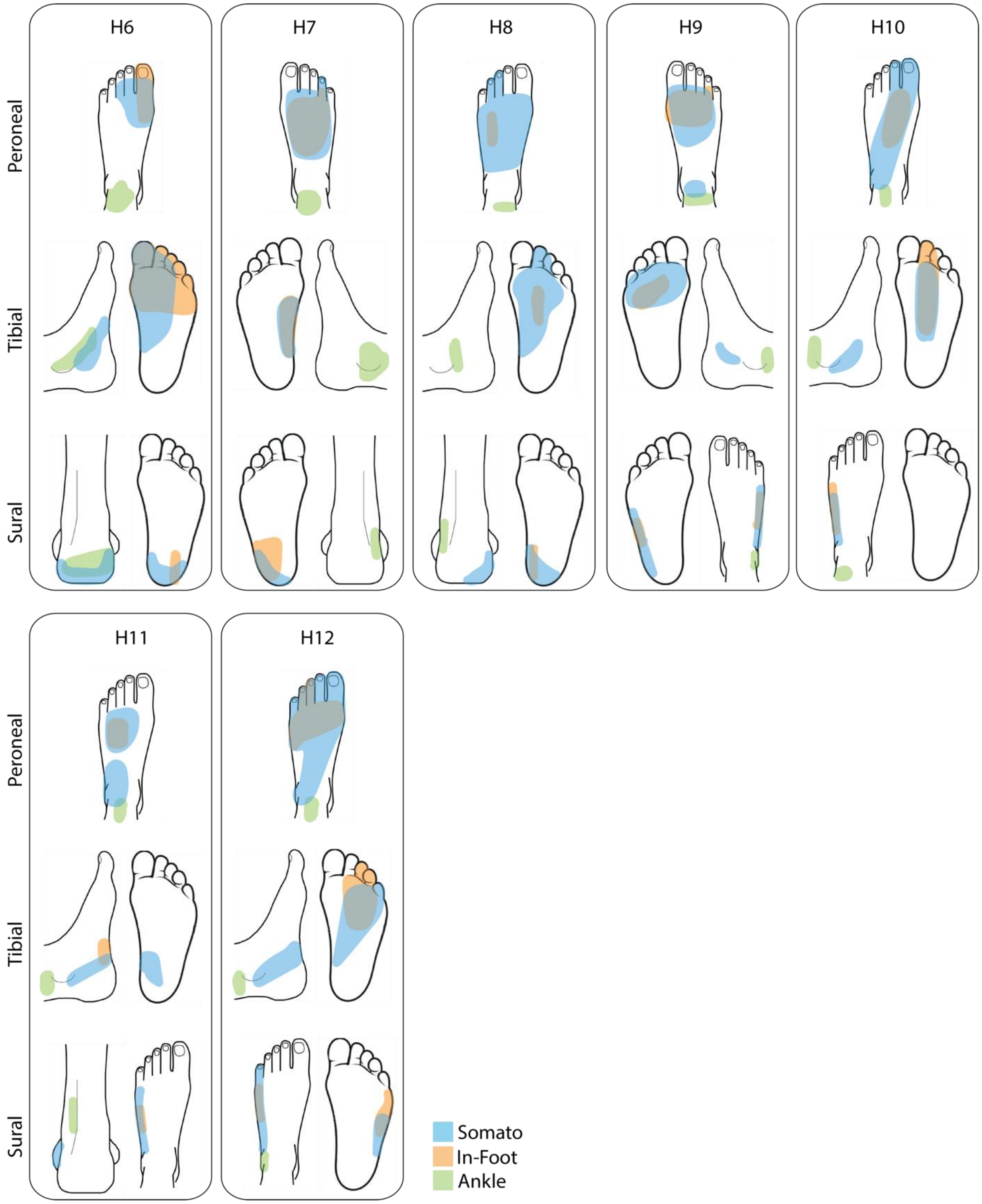


**Fig. S7 Calibration and areas of sensation of somatotopic, in-foot and on-ankle neurostimulation for fMRI experiments.** H: healthy participants. S: PN participant. For each participant three different locations (sural, tibial and peroneal) in three different conditions (somatotopic, in-foot, on-ankle) are stimulated. The three different conditions are overlapped for each location.

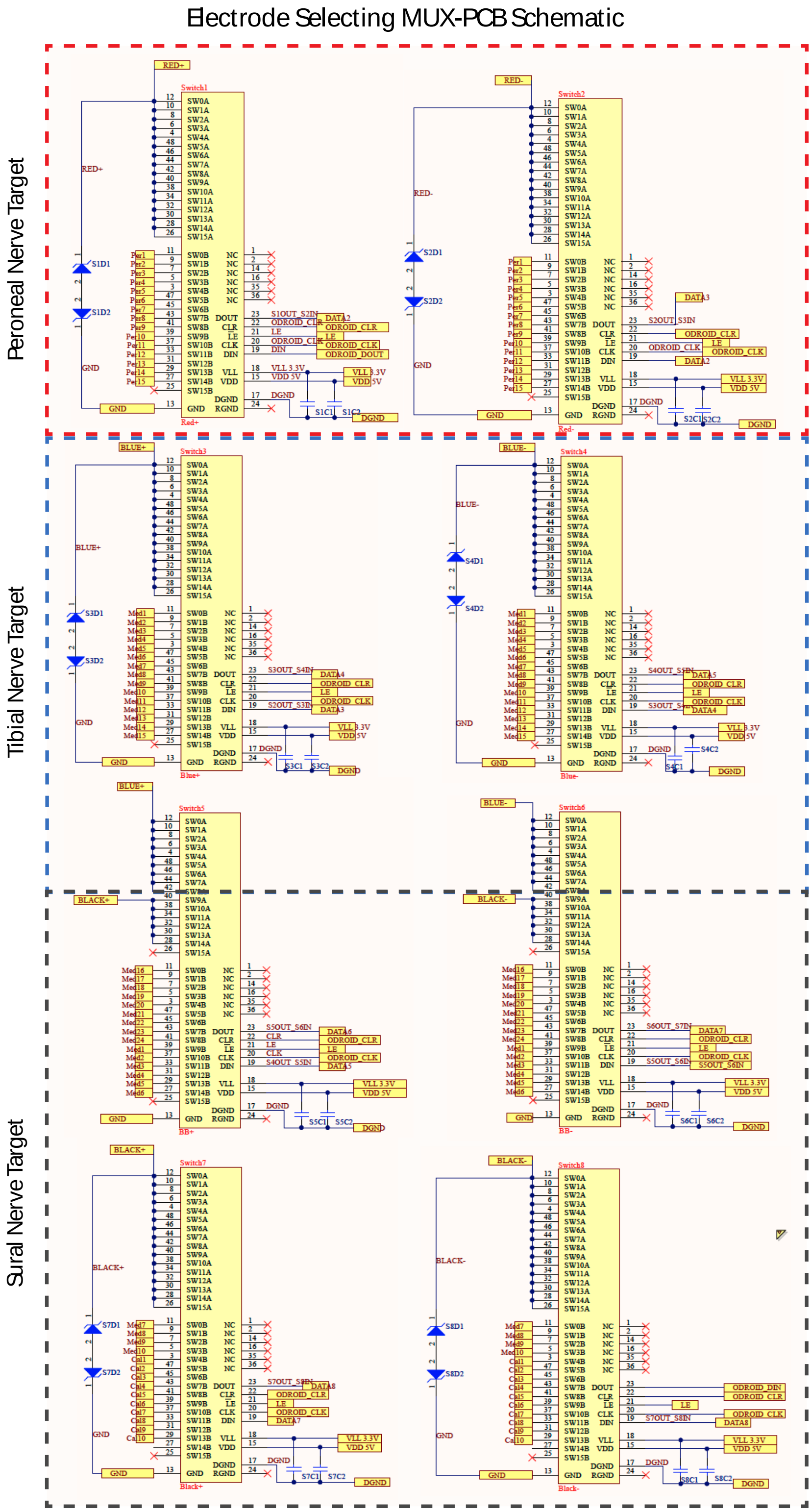


**Fig. S8. Electrode Selecting MUX-PCB Schematic.** Schematic of MUX PCBs that select which electrodes are active in the array. Colour labels (i.e. RED, BLUE, BLACK) refer to stimulator channels.

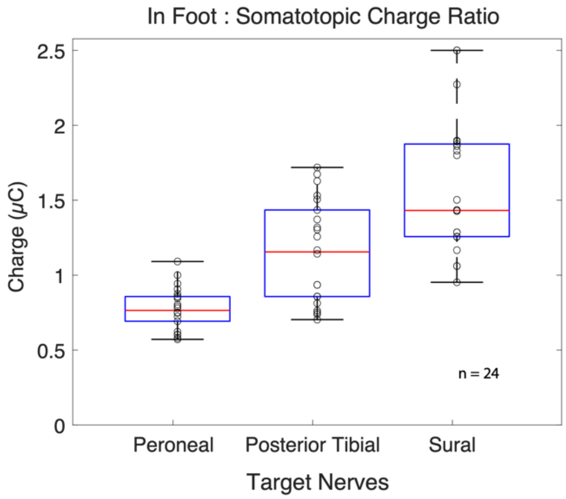

**Fig. S9. Healthy Charge Ratio for Different Target Nerves.** Charge ratio between the In Foot and Somatotopic conditions in the three target nerves.
